# Atlas of glomerular disease-specific genetic effects on blood transcriptome

**DOI:** 10.64898/2026.06.22.26356281

**Authors:** Lili Liu, Chen Wang, Oleksandr Kravets, Damian Fermin, Felix Eichinger, Francesca Zanoni, Atlas Khan, Jun Y. Zhang, Yan Ouyang, Qin Li, Patrick Hamilton, Philip A. Kalra, Rajkumar Chinnadurai, Kimberly Reidy, Jeffrey Kopp, Krzysztof Mucha, Cathy Smith, Abigail R. Smith, Michelle Mcnulty, Sean Eddy, Viji Nair, Margaret E. Helmuth, Tetyana L. Vasylyeva, William E. Smoyer, Celine Berthier, Rulan Parekh, Scott E. Wenderfer, Elizabeth Onugha, Tess Martin, Ksenia Sokolova, Rachel S.G. Sealfon, Chandra L. Theesfeld, Afshin Parsa, Rasheed Gbadegesin, Matthew Sampson, Simone Sanna-Cherchi, Olga G. Troyanskaya, Dirk S. Paul, Slave Petrovski, David Goldstein, Laura Heyns Mariani, Ali Gharavi, Columbia Genomics Consortium, NEPTUNE Consortium, CureGN Consortium, Matthias Kretzler, Krzysztof Kiryluk

**Author notes:** **Co-corresponding Authors:** Matthias Kretzler, M.D., Warner-Lambert/Parke-Davis Professor, Internal Medicine-Nephrology, Professor, Computational Medicine & Bioinformatics, University of Michigan, 1301 Catherine St, Med Sci 1, B-Wing, 3rd Floor, Room 3B3090, Ann Arbor, MI, 48109-5624, Krzysztof Kiryluk, M.D., M.S., Jay Meltzer Professor of Nephrology & Hypertension, Chief, Division of Nephrology, Columbia University, William Black Medical Research Building, 650 West 168th Street, 8th Floor, BB8-801F, New York, NY 10032. equal contributions (Lili Liu, Chen Wang).

## Abstract

IgA nephropathy (IgAN), IgA vasculitis (IgAV), focal segmental glomerulosclerosis (FSGS), membranous nephropathy (MN), and minimal change disease (MCD) account for the majority of idiopathic glomerulonephropathies (GN). These disorders involve immune system dysregulation and have a complex genetic architecture. Currently, there are no adequately powered blood transcriptomic datasets coupled to genetic data from patients with GN that can delineate disease-context specific genetic effects on the blood immune cell transcriptome. We performed whole genome sequencing coupled with bulk blood transcriptome sequencing on 1,822 participants from the CureGN study, a prospective cohort of participants with a kidney biopsy diagnosis of primary GN. We generated disease-context specific transcriptome-wide maps of gene expression QTL (eQTL), splicing QTL (sQTL), and double strand RNA-editing QTL (edQTL) for FSGS (N=447), IgAN (N=403), IgAV (N=123), MCD (N=408), and MN (N=441), as well as cross-disease maps for all 1,822 participants. Our QTL mapping identified 16,068 eGenes, 4,644 sGenes and 4,611 edQTLs with an FDR<0.05 in at least one GN type. Approximately 5-10% of the QTL signals were unique to a specific GN type, while ∼90% were shared between at least two conditions. Colocalization analysis demonstrated that ∼80% of shared eGenes between traits also shared the same causal variants, whereas ∼2% had distinct causal variants, suggesting context-specific regulatory effects. Cross-phenotype QTL mapping uncovered 6,466 eGenes, 2,705 sGenes and 5,321 edQTLs not previously detected in GTEx. Age, eGFR, and proteinuria-interaction QTL analyses identified hundreds of loci modified by age or disease severity. Lastly, integrative analyses with GWAS nominated new candidate genes for each of the five GN types under study. In summary, we generated comprehensive maps of GN-context-specific genetic effects on the blood transcriptome, providing a powerful new resource for integrative gene discovery studies of primary GN.

## Introduction

Primary glomerulonephropathies (GN) including IgA nephropathy (IgAN), IgA vasculitis (IgAV), focal segmental glomerulosclerosis (FSGS), membranous nephropathy (MN), and minimal change disease (MCD) represent important causes of kidney failure worldwide^1^. These disorders have complex genetic architecture, with genome-wide association studies (GWAS) having identified over 30 risk loci for IgAN^2–6^, 12 for MCD^7^, 4 for MN^8,9^, and 3 for IgAV^10–13^. Yet, translating GWAS findings into molecular mechanisms remains challenging, and is limited by the lack of genotype-informed regulatory maps derived from large and well-phenotyped cohorts of GN patients.

Immune dysregulation is central to the pathogenesis of GN, but the type of immune trigger, the molecular effector pathways, and the resulting kidney lesions differ substantially across diagnoses. IgAN and its related systemic disorder IgAV are prototypical immune complex-mediated diseases. In both of these conditions, aberrant mucosal immune responses lead to increased production of galactose-deficient IgA1 and anti-glycan antibodies, promoting formation of pathogenic immune complexes in the blood that deposit in the glomerular mesangium and lead to kidney injury^14^. MN exemplifies organ-specific autoimmune disease of the glomerular filtration barrier, where autoantibodies, most commonly directed against podocyte antigen PLA2R, form subepithelial immune deposits along the glomerular basement membrane^15^. There is also increasing evidence that a large fraction of MCD and FSGS represents an immune-mediated podocytopathy driven by circulating factors, such as anti-nephrin antibodies, causing glomerular injury^16^. A GN context-specific quantitative trait locus (QTL) framework is therefore well-suited to distinguish broadly shared and subtype-specific genetic regulatory mechanisms on gene expression, splicing and editing in circulating blood immune cells^17^.

In this study, we leveraged the CureGN cohort to generate a large genomic resource for five major GN types spanning across lifespan and diverse ancestries. We performed whole-genome sequencing (WGS) on 4,307 individuals with primary GN and 3,845 healthy controls, alongside whole-blood RNA sequencing in 1,822 GN cases with matched WGS data. For each GN subtype, we conducted WGS-based genome-wide case-control association analyses to identify common and rare variant associations. To define how genetic risk translates to gene dysregulation in GN, we mapped multiple layers of quantitative trait loci (QTLs) in the context of individual GNs, including expression QTLs (eQTLs), splicing QTLs (sQTLs), and A-to-I RNA-editing QTLs (edQTLs). We incorporated key covariates (age, sex, genetic ancestry, latent technical factors) and evaluated the impact of blood cell-type composition via computational deconvolution, enabling refinement of signals that may be masked or confounded by cellular heterogeneity. We further assessed allele-specific expression as an orthogonal line of evidence for cis-regulatory effects. Sharing and specificity of the genetic regulatory architecture was compared across GN types, and integration of QTLs with GWAS results prioritized new candidate molecular mediators for several established risk loci.

## Results

### Study Design

We conducted WGS at 30x depth on 4,307 individuals with primary biopsy-diagnosed GN, including the CureGN study^18^, and 3,845 healthy controls of diverse ancestries (**Figure S1**). The WGS cohort included 649 patients with FSGS, 1,513 with IgAN, 377 with IgAV, 1,023 with MN, and 745 with MCD. Whole blood RNA sequencing was also performed for 2,014 CureGN participants, including 1,822 patients with WGS data (447 FSGS, 403 IgAN, 123 IgAV, 408 MCD, and 441 MN cases). The study flowchart is depicted in **Figure 1**. First, we conducted genome-wide common and rare variant association analyses aiming to replicate known disease associations using newly generated WGS data. Next, we investigated the regulatory genetic architecture by GN-specific e/s/ed-QTL mapping and analyses of patterns of distinct and shared QTLs across GN types. We then performed integration of GWAS and GN-context-specific QTLs to better understand regulatory mechanisms underlying known GWAS loci. We also performed immune cell-type-specific interaction QTL mapping to define a set of QTL signals specific to each immune cell type and GN type. Interaction QTL analyses for age, sex, eGFR, and proteinuria were performed to identify age, sex, and disease-severity dependent QTL effects. Lastly, we performed cross-disease QTL mapping with additional adjustment for disease phenotypes. Our study provides a comprehensive atlas of GN-specific genetic effects on blood gene expression, splicing, and editing that can be used to accelerate genetic discoveries in GN.

**Figure 1.**
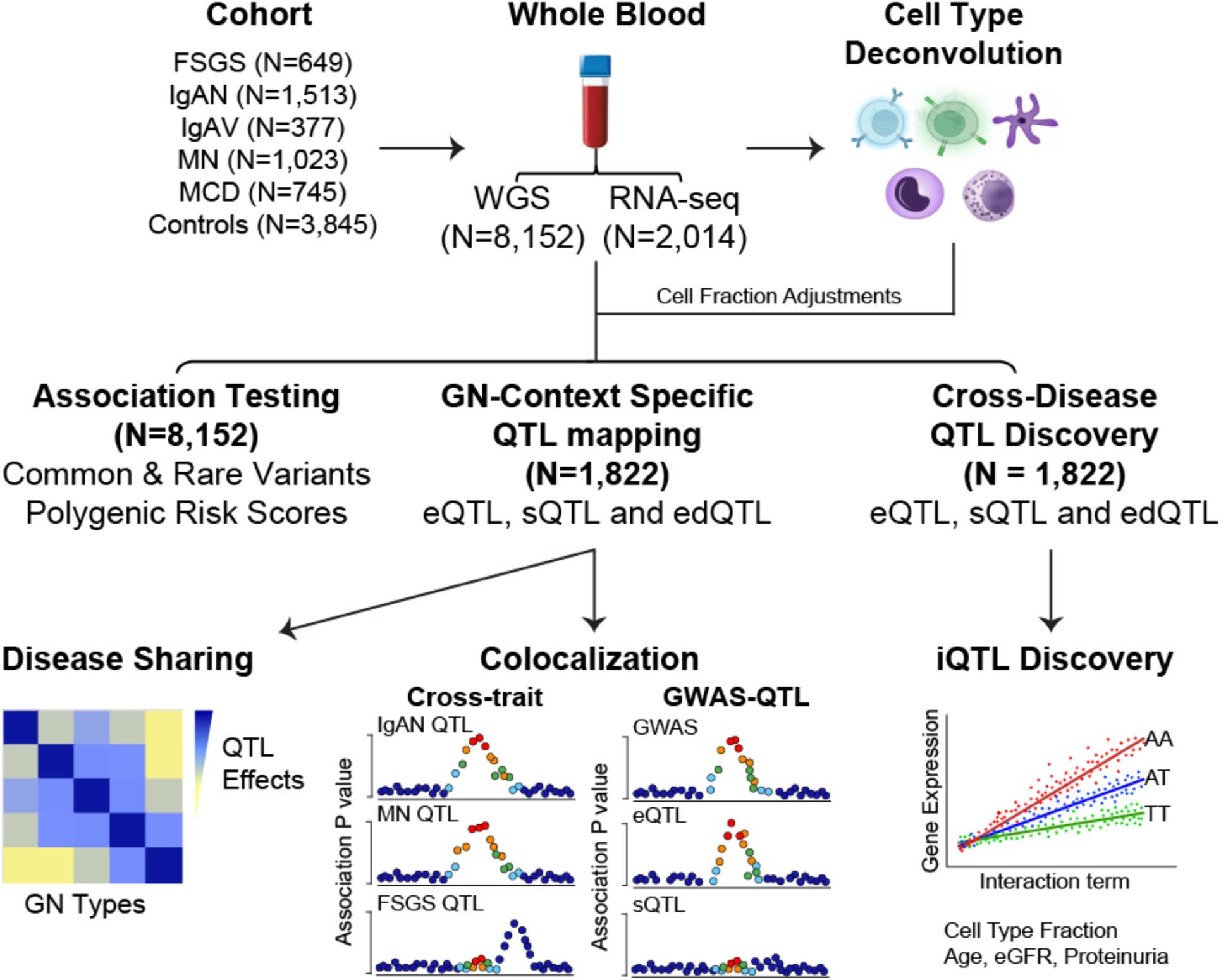
Study Design. Whole genome sequencing (WGS) was performed on 4,307 individuals with a kidney biopsy diagnosis of primary glomerulonephropathy (GN) and 3,845 healthy controls of diverse ancestries. Whole blood RNA sequencing was conducted for 1,822 GN patients with matched WGS data. Genome-wide case-control association analyses were carried out for each GN subtype to identify common variant associations, and gene- and gene-set-level collapsing analyses assessed rare variant burden. Polygenic risk scores were calculated and validated for each GN type. Transcriptome-wide mapping of context-specific quantitative trait loci (QTLs), including eQTLs, sQTLs, and edQTLs, was performed for each GN type, adjusting for deconvoluted cell type fractions. QTL sharing and specificity were examined, revealing genes regulated by shared or distinct causal variants by GN type. Integration of GWAS and QTL maps identified new GN-specific regulatory mechanisms and refined known loci. Cross-disease QTL analyses were performed to maximize discovery power and interaction QTL mapping further characterized genetic effects on gene regulation related to cell type, aging and disease severity. Additional abbreviations: FSGS: focal segmental glomerulosclerosis; IgAN: IgA nephropathy; IgAV: IgA vasculitis; MN: membranous nephropathy; MCD: minimal change disease.

### Common and rare variants association testing for GN subtypes

After joint calling and harmonization of WGS dataset for all 8,152 individuals, we performed genome-wide single variant genetic association analyses stratified by ancestry for each GN type, followed by meta-analysis (**Methods, Figure S2**). Several known large-effect loci surpassed genome-wide significance thresholds (P<5×10^−8^), including the *HLA* region (IgAN^5^, MN^8,9^, MCD^7^, and IgAV^13^), *PLA2R1* in MN^8,9^, and *APOL1* in FSGS^19^, and the majority of previously published loci were replicated at a nominal significance level (P<0.05), all with concordant effects between our cohort and previously published studies (**Figure S3**). We also performed multiple rare variant collapsing analyses and aggregation tests for coding and non-coding variants, including gene-based tests, gene-set analyses, regulatory region-based approaches, and sliding window scans (**Methods**). In the rare variant gene-set collapsing analyses, we identified significant associations between FSGS and two Human Phenotype Ontology (HPO)^20^ gene sets: minimal change glomerulonephritis (OR 2.00, P=3.9×10^−7^) and foamy urine (OR 1.95, P=2.2×10^−6^). Across all other complementary strategies, we did not identify new associations reaching genome- or exome-wide significance, suggesting that larger sample sizes will be required to further characterize the contribution of rare variants to GN susceptibility.

### Polygenic risk scores for GN subtypes

Using WGS data, we next validated the performance of the previously published polygenic risk scores (PRS) for GN and characterized their performance against both healthy and disease controls (all other unrelated GN types, **Figure S4**). For IgAN, the best performing PRS was a genome-wide score^5^ (AUC 0.681 against disease controls and 0.684 against healthy controls), which outperformed the simpler 30-SNP score. In contrast, for MN, the previously reported simple 6-SNP GRS^9^ showed stronger discrimination than the genome-wide polygenic score (AUC = 0.736 vs 0.680 against disease controls; 0.730 vs 0.686 against healthy controls). The steroid sensitive nephrotic syndrome PRS^7^ discriminated MCD from other GN types with AUC 0.648 and from healthy controls with AUC 0.637. Across all conditions, the AUC estimates were highly consistent between disease and healthy control comparisons, suggesting that the risk prediction models had reproducible performance even when applied to patients with other forms of GN.

We next explored, within each disease-specific cohort, the associations of best-performing PRS models with clinical disease features. The steroid sensitive nephrotic syndrome PRS was strongly associated with younger age at diagnosis (Beta: −4.5 years per PRS standard deviation, 95%CI: −6.1--2.9, P=3.6×10^−7^), replicating previous findings^7^. Moreover, MCD patients with high PRS were more likely to have a frequently relapsing course (OR: 1.28, 95%CI: 1.08-1.53, P=5.0×10^−3^). IgAN patients with high IgAN PRS were less likely to present with infections at disease onset (OR 0.77, 95%CI: 0.60-0.99, P=4.7×10^−2^). In MN, the PRS was associated with histologic PLA2R positivity (OR 1.88, 95%CI: 1.32-2.67, P=5.4×10^−4^), C3 staining intensity on immunofluorescence (OR 1.13, 95%CI: 1.04-1.22, P=7.0×10^−3^), and increased treatment resistance (OR 1.84, 95%CI: 1.35-2.52, P=2.3×10^−4^). The full set of PRS clinical correlations is provided in **Table S1**.

### Expression QTL mapping

We next explored the landscape of genetic regulation of blood cell gene expression in the context of GN. We tested for associations between blood gene expression levels and the genetic variants located within 1 Mb of the target gene’s start site (TSS), referred to as cis expression QTL mapping (cis-eQTL)^21^. The eQTL mapping was performed using a linear regression model, controlling for age, sex, genetic ancestry, and latent factors in the expression data^22^ (**Methods**). We further adjusted for the estimated cell fractions (CF) of five major blood cell types estimated by *in silico* computational deconvolution of bulk blood RNA-seq data to account for variability in cell composition across individuals^23–25^ (see **Methods** and **Figure S5**). At 5% false discovery rate, we identified a total of 16,068 genes with a significant cis-eQTL association referred as eGenes, in at least one GN phenotype with CF adjustment (**Figure 2a, Table S2-6**). Comparing CF-adjusted and unadjusted results revealed that 83-99.7% of eGenes were consistent between the two analyses (**Figure S6**). A total of 374 eGenes lost significance across all GN types, while we observed 339 new eGenes after accounting for CFs (**Table S7**). Additionally, we performed interaction eQTL (ieQTL) mapping across major blood cell types using computational estimates of cell type proportions to identify cell type-specific eQTL (see **Supplemental Results, Figure S7** and **Table S8-S12**).

**Figure 2.**
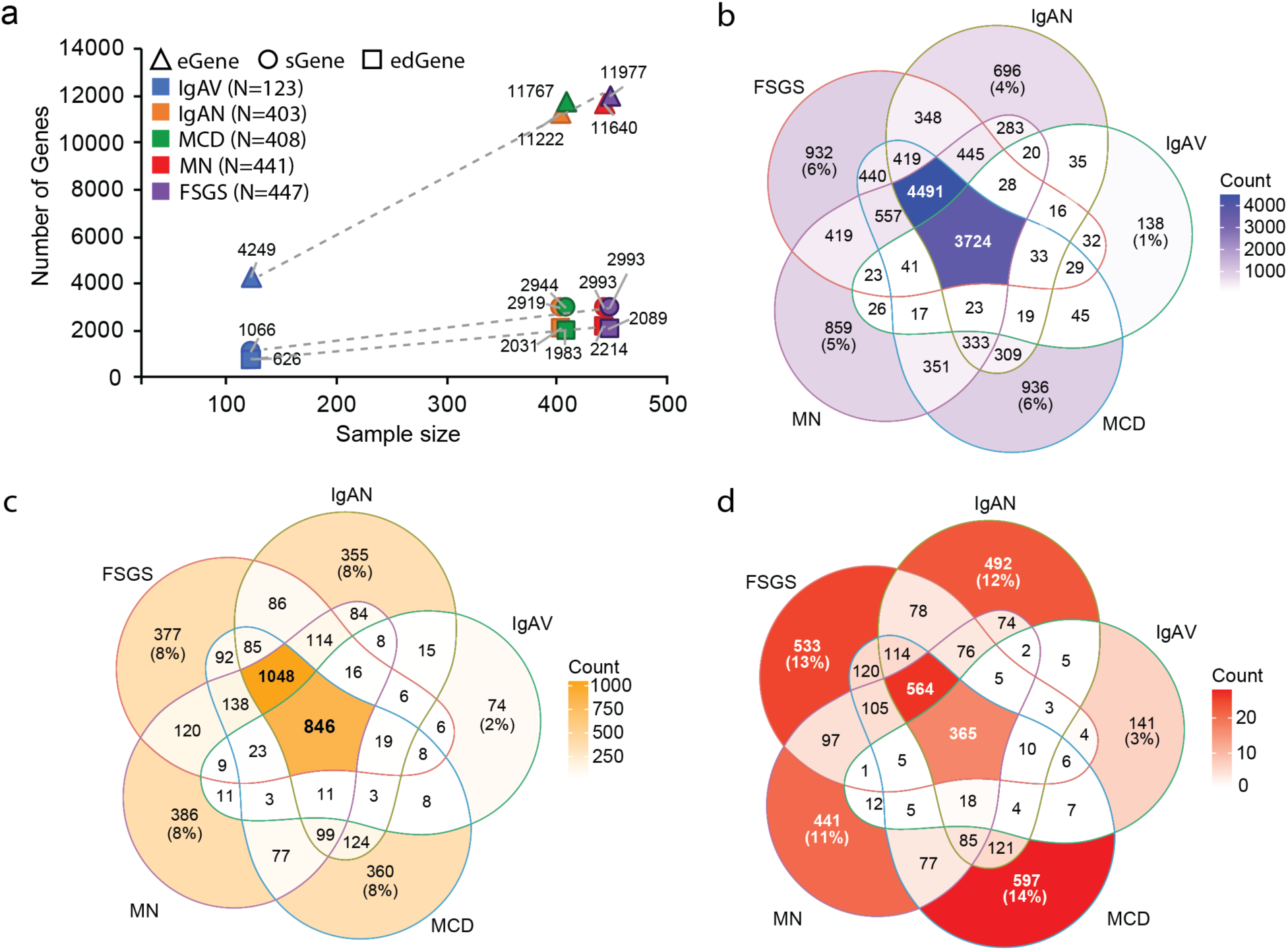
Discovery of disease context-specific QTLs in the CureGN cohort. (a) Number of eGenes (triangles), sGenes (circles), and edGenes (squares) identified for each GN subtype (IgAV, IgAN, MCD, MN, FSGS) in relation to sample size. (b) Venn diagram showing overlap of identified eGenes across GN subtypes. (c) Venn diagram illustrating overlap of sGenes. (d) Venn diagram depicting overlap of edGenes. Percentages indicate the proportion of GN subtype-specific genes. Counts in shaded regions represent the number of genes shared among two or more GN subtypes. Color gradients indicate the number of overlapping genes.

Consistent with previous studies^21^, the number of eGenes was positively associated with the sample size of each GN cohort (**Figure 2a**). Because gene expression often exhibits substantial cis allelic heterogeneity, whereby a single gene can be regulated by multiple independent QTLs^21^, we also mapped independent cis-eQTLs for each phenotype using stepwise regression^21^. We confirmed widespread allelic heterogeneity, with greater than 40% of eGenes having more than one independent cis-eQTL in IgAN, MCD, MN and FSGS (each with sample size > 400) (**Figure S8a, Table S13-17**). In IgAV (smallest cohort), we detected 13.6% of eGenes with more than one independent eQTL (**Figure S8a; Table S17**). Comparing genome-wide genetic effects on gene expression across major GN types revealed shared and distinct patterns of genetic regulation (**Figure S8b**). Examining the overlap of eGenes across different GN types, we observed that 5-6% of eGenes were significant only in a single GN type (**Figure 2b**), except for IgAV (smallest cohort), which had ∼1% of IgAV-specific eGenes.

Allele-specific expression (ASE) analysis provides an orthogonal complement to cis-eQTL mapping by leveraging reads overlapping heterozygous sites to quantify allelic imbalance within the same individual. Therefore, we used transcriptome-wide ASE analysis to validate our cis-eQTL signals (see **Supplemental Results** and **Figure S9**). Large proportions (39-52%) of cis-eQTL signals were confirmed with the ASE analysis using our novel direction-based approach to define significant ASE associations (see **Methods**). We also found strong correlation between the cis-eQTL slope estimates and ASE effect sizes among signals significant in both cis-eQTL and ASE analyses (Spearman’s ρ = 0.88-0.92).

### Splicing QTL mapping

We further investigated the impact of genetic variation on splicing of protein-coding and long intergenic noncoding RNA (lincRNA) genes. For each GN type, we mapped sQTLs in cis with intron excision ratios from LeafCutter^26^, controlling for age, sex, genetic ancestry, and latent factors in the splicing levels as well as estimated cell type fractions. In the analysis of 35,097 splicing events for 12,840 genes, we discovered 4,611 sGenes (36%) with a significant cis-sQTL in at least one GN type (**Figure 2a, Table S18-22**). Comparison of CF-adjusted and CF-unadjusted sQTL mapping showed that about 93% of significant sGenes were consistent between the two analyses, and 150 additional sGenes were identified across GN forms after accounting for CFs (**Figure S10, Table S23**).

Similar to eQTL analyses, we observed positive correlation between the number of sGenes and sample size across GN cohorts (**Figure 2a**), ranging from 2,993 sGenes discovered for FSGS (largest cohort) to 1,066 discovered for IgAV (smallest cohort). Over 90% of these sGenes were shared across at least two phenotypes, while about 8% were condition-specific (**Figure 2c, Table S18-22**). Notably, in IgAV, the cohort with the smallest sample size, we identified only 74 IgAV-specific sGenes, but they were significantly enriched in adaptive immune responses (FDR=1.1×10^−3^) and B cell-mediated immunity (FDR=3.3×10^−2^), suggesting that disease context-specific regulation of splicing was related to systemic immune activation in IgAV.

### Double-strand RNA-editing QTL mapping

Previous studies have demonstrated contributions of RNA editing to human immune-mediated disorders^27^. Particularly, the adenosine-to-inosine (A-to-I) editing, one of the most abundant RNA modifications, suppresses double-strand RNA (dsRNA) sensing mediated by MDA5, a cytosolic sensor of ‘non-self’ dsRNA^28–31^. Moreover, A-to-I RNA editing has emerged as another potential molecular mechanism connecting GWAS-identified risk alleles to disease susceptibility^32,33^. To further investigate which variants may contribute to differences in RNA editing in patients with GN, we performed A-to-I editing QTL (edQTL) in each GN type (**Methods**). We first quantified RNA editing levels, defined as the fraction of edited (‘G’) transcripts relative to total (‘A’ + ‘G’) transcripts, at a single-nucleotide (“site”) resolution across more than 2.8 million annotated RNA editing sites. This yielded reliable editing-level estimates for 21,547-25,454 sites across 1,530-1,674 edited genes (edGenes) per GN type (**Figure S11a; Methods**). Interestingly, despite smallest sample size, IgAV exhibited a significantly higher number of identified editing sites compared with other GNs (**Figure S11a**). Most genes showed limited editing with <10 identified editing sites (63%), whereas approximately 7% were highly edited, with more than 100 identified editing sites across CureGN participants (**Figure S11b**). The *SPN* gene (sialophorin, also known as CD43) was the most edited gene, with an average of 325 detected editing sites across GN forms. Consistent with previous studies^27,29^, a large proportion of editing sites (57%) were located in UTRs or exonic regions (**Figure S11c**), unlike the eQTLs (enriched near the transcription start site) or sQTLs (enriched near splice junctions)^21^.

Next, we mapped edQTLs for each GN type, controlling for age, sex, genetic ancestry, and latent factors in editing levels. Of all tested editing sites, we identified significant edQTLs for 4,611 sites with FDR<0.05 in at least one GN phenotype (**Table S24-S28**). Similarly, the number of edSites per phenotype was dependent on sample size and overall editing levels (**Figure 2a**). Among the significant edQTLs, the majority (over 80%) were shared across at least two GN forms, whereas about 3-14% of the edQTLs were disease-context specific (**Figure 2d, Table S24-S28**). Interestingly, *CTSS* (encoding Cathepsin S, which participates in the degradation of antigenic proteins to peptides for presentation on MHC class II molecules) was a significant edGene with seven significant edQTL effects at seven different editing sites identified in FSGS and two in MCD, while no significant edQTLs were observed in other GN types. A previous study demonstrated that highly edited *CTSS* enables the recruitment of the stabilizing RNA-binding protein human antigen R to the 3′ UTR of the *CTSS* transcript, thereby controlling *CTSS* mRNA stability and expression^34^. Mechanistic studies are needed to understand the consequences of the altered genetic regulation of Cathepsin S in the context of FSGS and MCD.

### Genetic regulatory effects across glomerular disorders

The context-specific QTL mapping across different GN forms indicated that the majority of the genetic regulation on gene expression, splicing and RNA-editing are shared across at least two phenotypes. To investigate whether shared QTLs across traits are mediated by the same underlying genetic regulatory mechanisms, we further performed colocalization for each eGene shared between traits. We observed that the majority (∼80%) of shared eGenes between traits had >80% posterior probability of sharing a causal variant (PP4>80%, **Figure 3a, Table S29**). However, ∼2% of shared eGenes had >80% posterior probability of different causal variants between traits (PP3>80%), suggesting distinct genetic regulatory mechanisms of the same gene under a specific disease context (**Table S29**).

**Figure 3.**
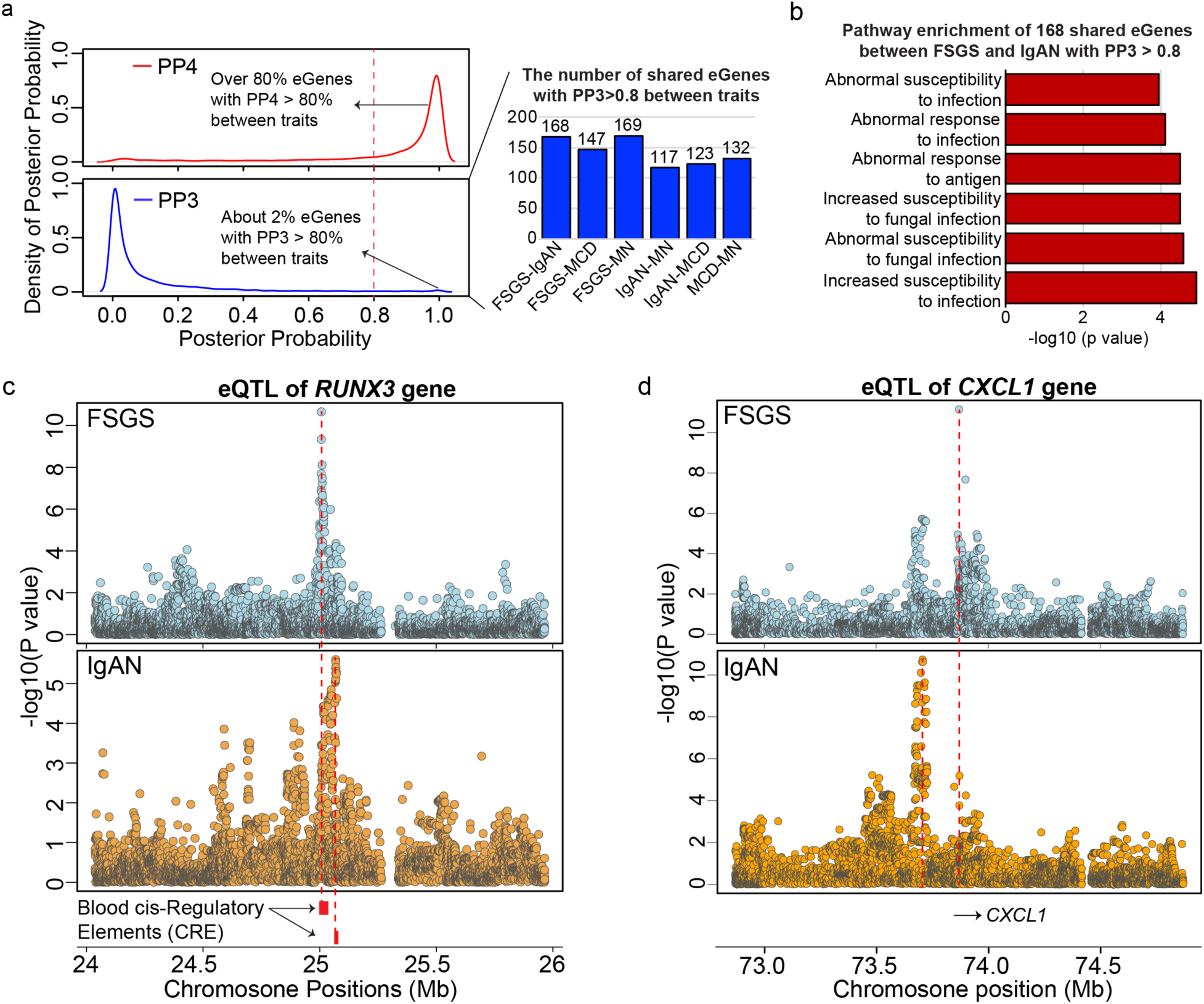
Shared and distinct genetic regulation on gene expression across GN forms. (a) Posterior probability distributions for eGene sharing between GN subtypes: PP4 (strong evidence for shared eQTLs) shows over 80% sharing between traits, while PP3 (distinct but colocalized signals) shows ∼2% sharing. Bar graph indicates the number of shared eGenes (PP3 > 0.8) between pairs of GN subtypes.(b) Pathway enrichment analysis of 168 shared eGenes between FSGS and IgAN with PP3 > 0.8 highlights pathways related to abnormal susceptibility and response to infection (c) Distinct patterns of non-colocalizing regional associations for eQTLs of the *RUNX3* gene in FSGS and IgAN patients, with lead SNPs intersecting distinct cis-regulatory elements (CRE) in each condition.(d) Distinct and non-colocalizing regional association plots for eQTLs of the *CXCL1* gene in FSGS and IgAN, providing another example of trait-specific genetic regulation of the same gene. Dashed red lines denote the lead eQTL variant position in each panel.

For example, there were 169 eGenes overlapping between FSGS and IgAN that had a PP3 greater than 80% (**Table S30**), and these genes were significantly enriched in multiple immune and infection-related phenotypes, including “abnormal susceptibility and response to infection”, “abnormal response to antigen”, and “increased susceptibility to fungal infection” (**Figure 3b**). This could be explained by the role of infections in susceptibility to IgAN, but not necessarily FSGS. As an example, the *RUNX3* gene (encoding RUNX Family Transcription Factor 3) previously implicated by GWAS in the regulation of serum IgA levels^35^ and IgG glycosylation^36,37^, was a significant eGene in both FSGS and IgAN cohorts (**Figure 3c**). However, the lead SNPs for each disease-specific eQTL signal were separated by a 60-kb distance and had a distinct pattern of reginal association with colocalization PP3 of 80%, consistent with distinct causal variants (**Figure 3c**). Each lead eQTL SNP intersects a separate blood cis-regulatory element (CRE)^38^, supporting activation of distinct genetic regulatory mechanisms underlying blood *RUNX3* expression under specific disease context.

### Cross-disease QTL discovery

We next conducted cross-disease QTL mapping across all five CureGN conditions (N=1,822) with additional adjustment for GN type. We detected 17,210 genes with an eQTL, 5,145 genes with an sQTL and 7,175 genes with an edQTL at 5% false discovery rate (**Figure 4a-c; Table S31-33**). Of these, 6,466 eGenes, 2,705 sGenes and 5,321 edGenes were not previously detected in the GTEx blood tissue (**Figure 4a-c**). The pathway enrichment analysis indicated that the newly discovered eGenes in the CureGN cohort were significantly enriched in multiple immune related pathways, including cellular responses to stimuli, viral infection pathways, and cytokine signaling in immune system (**Figure 4a**). The newly discovered sGenes were significantly enriched in adaptive immune system and T cell activation (**Figure 4b**), while the newly discovered edGenes were significantly enriched in innate immune system, neutrophil degranulation, and antigen activates B cell receptor (**Figure 4c**).

**Figure 4.**
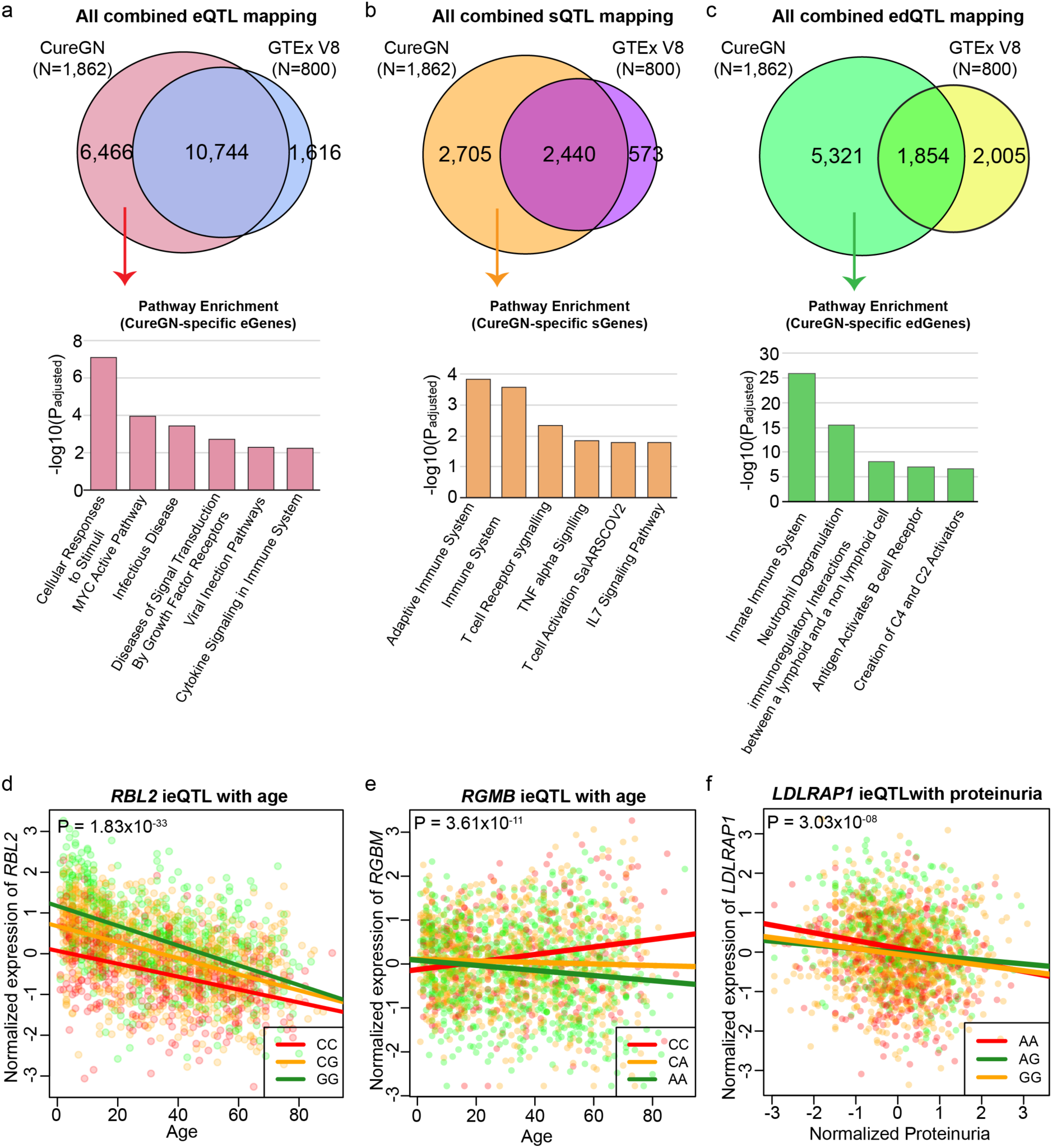
Cross-phenotype QTL and interaction QTL mapping in 1,822 CureGN participants. **(a-c)** Venn diagrams showing overlap and unique QTL findings between the CureGN cohort and GTEx V8 blood samples: **(a)** combined eQTL mapping, **(b)** combined sQTL mapping, and **(c)** combined edQTL mapping Unique QTLs identified in CureGN are significantly enriched in multiple immune-related pathway enrichment. **(d-f)** Scatter plots illustrating the effects of interaction QTLs on gene expression (ieQTL) with age and proteinuria: **(d)** *RBL2* ieQTL with age. (e) *RGMB* ieQTL with age. (f) *LDLRAP1* ieQTL with proteinuria. Color coding denotes genotype groups in each plot.

### Age-related QTLs

Age is the risk factor for many common diseases, including kidney disorders^39–41^, and genotype-phenotype associations are often modified by age^42,43^. To understand how aging affects the genetic effects on blood transcriptome, we took advantage of the fact that the CureGN study recruited patients across the lifespan (age range 1-99 years). To model the impact of age, we performed genotype-by-age ieQTL mapping on blood transcriptome using all combined CureGN cohort (N=1,822), adjusting for GN type, sex, genetic ancestry, PEER factors, and estimated cell type fractions. We identified 232 age-dependent eQTLs based on FDR<0.05 (**Table S34**). To additionally account for potential confounding by estimated glomerular filtration rate (eGFR), we repeated the age-interaction eQTL analysis after controlling for eGFR. Of the 232 age-interaction eGenes, 159 remained significant (FDR <0.05) after eGFR adjustment (**Table S34**).

The strongest age-ieQTL was identified for the *RBL2* gene encoding p130, a well-established regulator of cell-cycle-dependent signaling and cellular senescence linked to ageing^44^. Downregulation of *RLB2* causes telomere shortening and premature senescence^45,46^. As demonstrated in **Figure 4d**, the blood expression of *RBL2* decreased significantly with age, and age significantly attenuated the genetic effect of rs9938788 on *RBL2* expression. While younger individuals exhibit divergent blood levels of *RBL2* expression by rs9938788 genotype, *RBL2* expression becomes progressively downregulated across genotype groups with older age, ultimately converging to comparably low levels irrespective of genotype.

Another age-related ieQTL at *RGMB (Repulsive Guidance Molecule BMP Co-Receptor B)* gene exhibits an opposite phenomenon, where the genetic effect is significantly enhanced with advanced age (**Figure 4e**). *RGMB* is expressed in multiple immune cell types, including antigen presenting cells (macrophages and dendritic cells)^47^. Increased expression of *RGMB* has been associated with immunosuppression, including suppression of anti-tumor immunity^47–50^. Individuals carrying the rs10648681-CC genotype tend to exhibit significantly higher *RGMB* expression at older ages compared to other genotype groups.

### Kidney disease severity-related QTLs

To further identify genetic effects on gene expression modified by the severity of kidney disease, we performed ieQTL mapping with eGFR and proteinuria, adjusting for age, sex, genetic ancestry, PEER factors, diagnosis as well as estimated cell type fractions. We identified 57 eQTLs with significant eGFR interactions based on FDR<0.05 (**Table S35**). Among them, 3 genes have been previously linked to eGFR levels by GWAS, including *RBL2*, *NBPF3,* and *SPG7*^51^.

We also identified 13 eQTLs with significant proteinuria interactions based on FDR<0.05 (**Table S36**). Among them, *BTAF1* and *RBL2* have been associated with eGFR levels in GWAS^51^. One interesting example is *LDLRAP1* gene encoding Low Density Lipoprotein (LDL) Receptor Adaptor Protein 1 essential for clearing blood cholesterol^52,53^; low expression of *LDLRAP1* has been associated with higher LDL levels and increased risk of cardiovascular disease^54^. Notably, the expression of this gene declined with increasing level of proteinuria, attenuating the eQTL effect of rs6661159 observed in non-proteinuric patients (**Figure 4f**). This effect of proteinuria on the genetic regulation of *LDLRAP1* expression could be potentially contributing to the hyperlipidemia commonly observed in patients with nephrotic syndrome.

### Integration of QTL maps with GWAS

GWAS have identified genetic susceptibility loci for multiple glomerular disorders, including IgAN^5^, IgAV^13^, MN^9^, and MCD^7^. Our atlas of GN-context specific QTL effects offers a unique resource to understand the functional consequences of these loci and to prioritize candidate causal genes from existing and future GWAS studies. To demonstrate its utility, we performed comprehensive colocalizations for each of the known GN loci with the corresponding e/s/edQTL signals within and across different GN types (**Figure 5a**). These analyses revealed multiple QTLs with strong support for sharing a causal variant with GWAS signals. Based on these results, we prioritized eGenes for 13 loci, sGenes for 11 loci and edQTL effects for 5 loci based on the conservative PP4>0.8.

**Figure 5.**
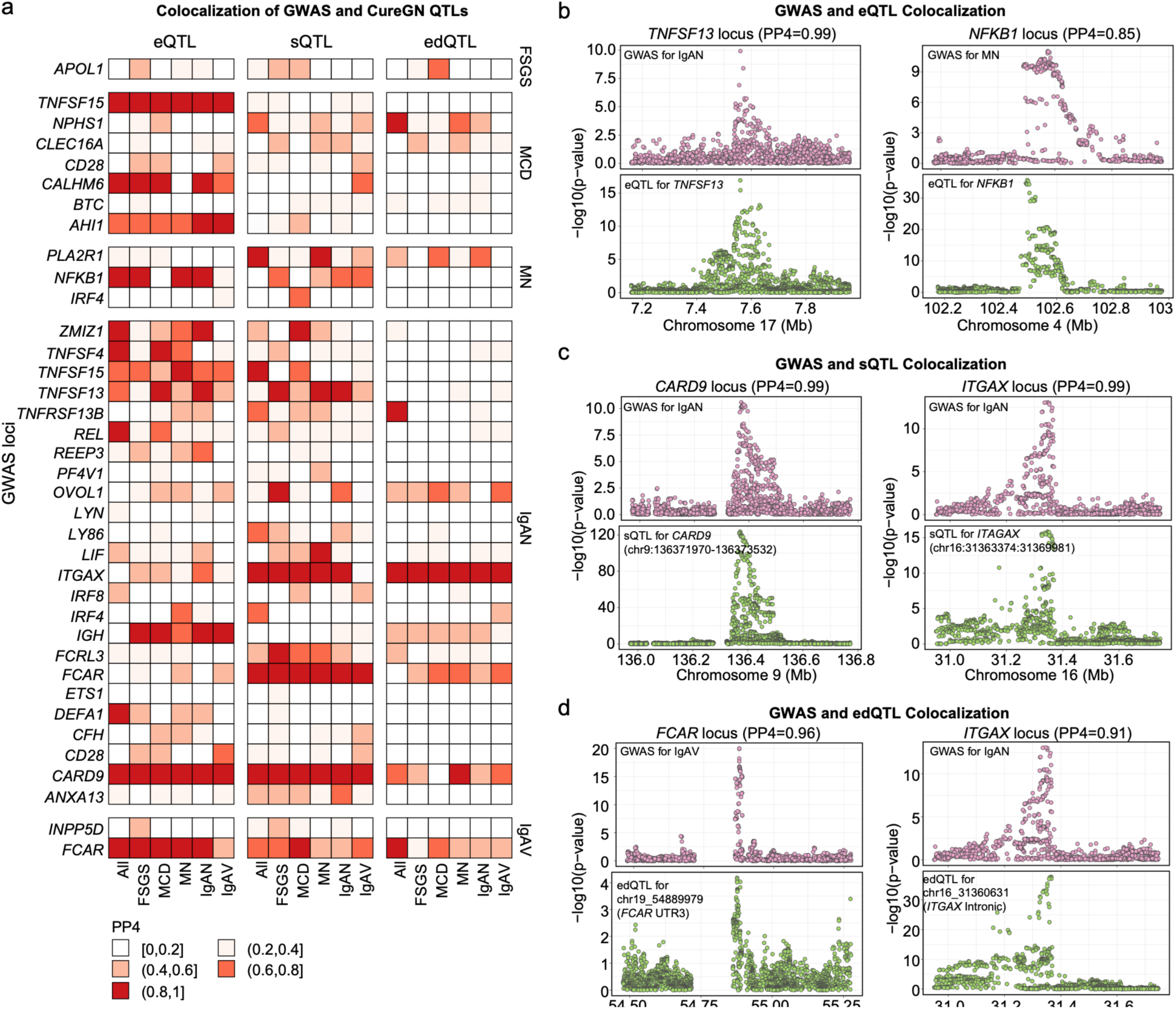
Colocalization of GN-associated non-HLA GWAS signals with CureGN blood e/s/edQTLs. **(a)** Heatmap summarizing posterior probability of sharing a causal variant (PP4) between GWAS loci for focal segmental glomerulosclerosis (FSGS), minimal change disease (MCD), membranous nephropathy (MN), IgA nephropathy (IgAN), and IgA vasculitis (IgAV) and their corresponding context-specific molecular QTLs (eQTL, sQTL, edQTL). Color scale denotes PP4 with higher values indicating stronger support for a shared causal variant. **(b)** Examples of regional association plots for colocalization of GWAS and eQTL signals at the *TNFSF13* locus for IgAN (PP4=0.99) and *NFKB1* locus for MN (PP4=0.85). **(c)** Regional association plots showing colocalization of GWAS and sQTL signals at the *CARD9* locus for IgAN (PP4=0.99) and *ITGAM/X* locus for IgAN (PP4=0.99). **(d)** Regional plots showing colocalization of GWAS and edQTL signals at the *FCAR* locus for IgAV (PP4=0.96) and *ITGAX* locus for IgAN (PP4=0.91). Genomic position is shown on the x-axis. Y-axis denotes -log10 (P value).

Here, we highlight several previously unreported colocalization signals between our disease-context specific QTLs and known GWAS loci for GN (**Figure 5b**; **Table S37**). For example, we detected a strong colocalization of the IgAN eQTL signal for *TNFSF13* with *TNFSF13* GWAS locus for IgAN (PP4=1.00), where the risk allele (rs3803800-A) was associated with increased blood *TNFSF13* expression (**Figure 6).** Another IgAN GWAS locus colocalized with the IgAN-specific eQTL for *ZMIZ1* (PP4=0.81), an immunoregulatory gene previously prioritized as a potential positional candidate^5^. Both *TNFSF13* and *ZMIZ1* GWAS loci were not colocalized with a much larger healthy blood eQTL reference dataset (N=31,684)^55^, highlighting the utility of our context-specific maps. We also independently validated the recently reported colocalization of the *FCAR* susceptibility locus for IgAV with *FCAR* eQTL, but not sQTL^13^. At the same time, the *FCAR* susceptibility locus for IgAN colocalized with *FCAR* sQTL, but not eQTL, suggesting distinct regulatory mechanisms in IgAN versus IgAV. For MN, we detected a new colocalization of the MN-specific eQTL for *NFKB1* with the *NFKB1* MN susceptibility locus (PP4=0.99). Lastly, for MCD we detected new colocalizations of *DSE* at *CALHM6* locus (PP4=0.99), *AHI1* at *AHI1* locus (PP4=1.00), and *TNFSF15* at *TNSF15* locus (PP4=0.99).

**Figure 6.**
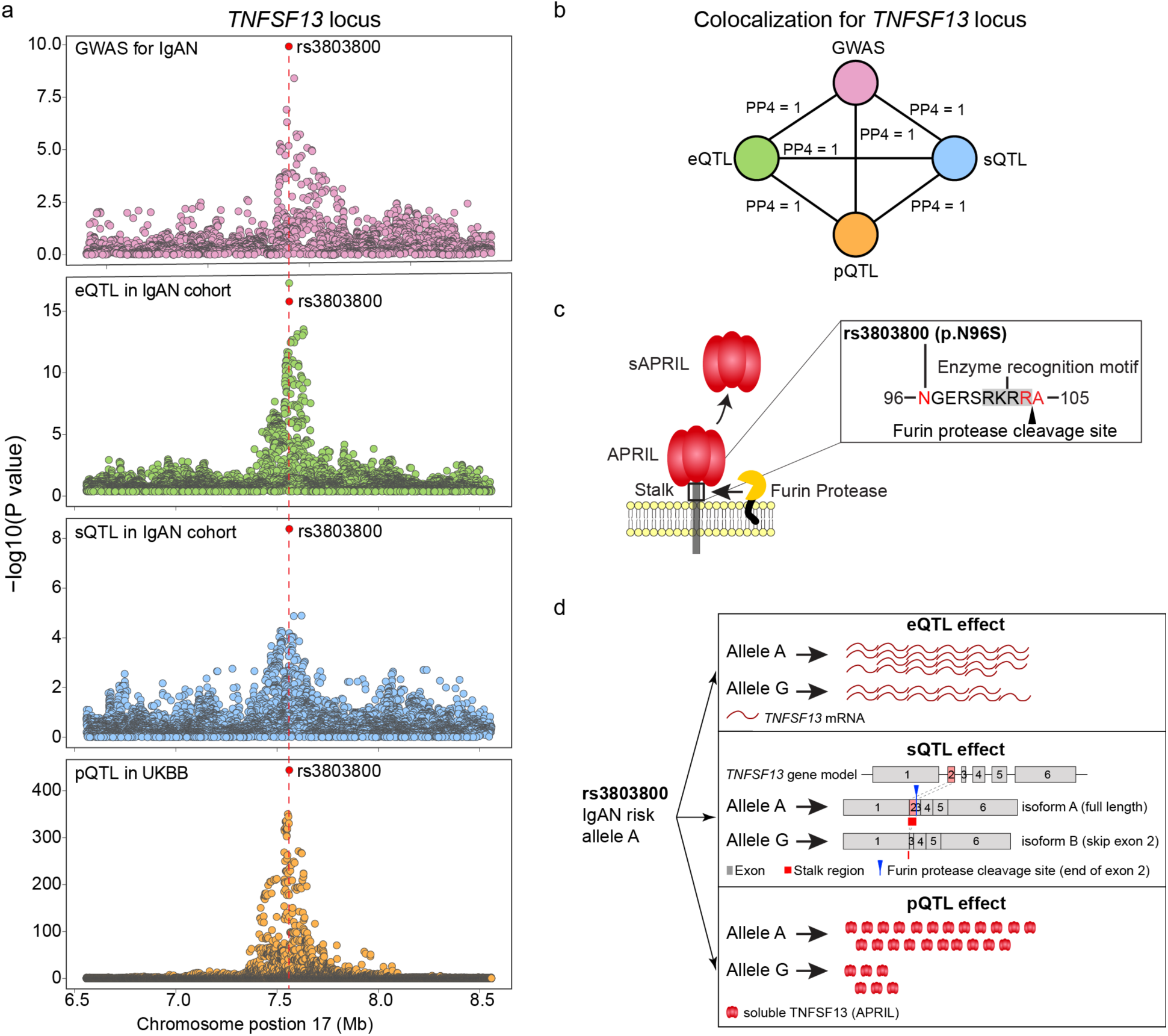
*TNFSF13* locus in IgA nephropathy (IgAN). **(a)** Regional plots of the *TNFSF13* locus in the latest IgAN GWAS (10,146 cases and 28,751 controls), expression QTL (eQTL) of *TNFSF13* in the IgAN cohort (N=403), splicing QTL (sQTL) for the chr17:7559297-7559846 splicing event in *TNFSF13* in the IgAN cohort (N=403), and plasma TNFSF13 (APRIL) protein QTL (pQTL) in the UK Biobank (UKBB, N= 54,219). **(b)** Co-localization between traits indicates a shared causal variant at IgAN *TNFSF13* locus with its eQTL, sQTL and pQTL effects. **(c)** The location of p.N96S variant (rs3803800) in relation to the cleavage site of the membrane bound APRIL. **(d)** The summary of QTL effects of rs3803800, the top IgAN GWAS SNP at the *TNFSF13* locus: the IgAN risk allele (rs3803800-A) is associated with higher blood levels of total *TNFSF13*, increased expression of the intact, fully functional *TNFSF13* transcript, and elevated levels of secreted soluble TNFSF13 (APRIL).

We also identified 14 novel splicing events colocalized with 11 GN-associated GWAS loci (**Table S37**), with selected examples shown in **Figure 5c**, including colocalizations between IgAN GWAS loci and sQTLs for *CARD9* (chr9:136371970-136373532; PP4=0.99) and *ITGAX* (chr16:31363374-31369981; PP4=0.99). The *CARD9* splicing event truncates exon 2, producing a shortened caspase recruitment domain (CARD). The IgAN protective allele is associated with truncation of the functional CARD domain, whereas the risk allele is linked to higher expression of the intact, active CARD9 isoform, indicating effects on both CARD9 expression and splicing. Notably, at the *TNFSF13* IgAN GWAS locus, we also observed strong colocalizations (PP4=1.00) across eQTL, sQTL (splicing event: chr17:7559297-7559846) and protein QTL (pQTL), consistent with a shared causal variant across all modalities (**Figure 6a and b**). The top GWAS SNP rs3803800 is a missense variant (p.Asn96Ser) located in the stalk region of membrane-bound TNFSF13 (also known as APRIL) and close to its cleavage site (**Figure 6c**). The *TNFSF13* splicing event (chr17:7559297-7559846) results in skipping of exon 2 of *TNFSF13*, which encodes the stalk region of membrane-bond TNFSF13 (**Figure 6c**). The IgAN protective allele (rs3803800-G) is associated with decreased *TNFSF13* mRNA expression in blood, increased exon 2 skipping, and reduced plasma levels of soluble APRIL. In contrast, the IgAN risk allele is associated with increased expression of the fully functional *TNFSF13* transcript in blood and significantly elevated plasma levels of APRIL (**Figure 6d**). APRIL is a powerful B-cell stimulating cytokine that increases IgA1 production and it is being targeted by two newly approved immunomodulatory drugs for IgAN^56,57^.

Additionally, we uncovered multiple novel dsRNA A-to-I editing events associated with GN risk, including the examples in **Figure 5d**. The IgAV *FCAR* locus colocalized with an editing event in the 3′ UTR of *FCAR* (chr19:54889979), and the risk allele was associated with decreased editing levels at this site. These findings suggested a pleiotropic effect of the IgAV risk allele associated with both upregulation of *FCAR* mRNA and downregulation of RNA editing at chr19:54889979. Similarly, we observed a pleiotropic molecular effect at the IgAN *ITGAX* risk locus characterized by altered *ITGAX* splicing (chr16:31363374:3136998) and reduced RNA editing at chr16_31360631 within the intronic region of *ITGAX*. **Table S37** summarizes top colocalization signals for all other GWAS loci.

## Discussion

In this study, we provided the largest WGS reference dataset for genetic studies of glomerular disorders comprised of 4,307 biopsy-diagnosed GN cases and 3,845 controls. Our genetic association analyses replicated known GWAS loci and validated previously published polygenic scores for various types of primary GN. By coupling genome with blood transcriptome sequencing in a subset of 1,822 cases, we also provided the first comprehensive disease context-specific atlas of genetic effects on blood transcriptome in primary GN. We demonstrated how this resource could be used effectively to prioritize GWAS candidate causal genes, and we made all results publicly available to enhance future gene discovery efforts for GN.

With the identification of 16,068 eGenes, 4,644 sGenes, and 4,611 A-to-I RNA editing sites with edQTLs, our study greatly expanded the catalog of genetic variants associated with transcriptomic effects in blood. Adjusting for estimated blood immune cell fractions uncovered additional 339 eGenes and 150 sGenes, indicating that heterogenous cell composition can mask some true cis-regulatory effects in patient cohorts profiled with bulk blood RNA-seq, and that cell fraction adjustment improves discovery rates.

Importantly, our analyses underscore the importance of disease context, revealing that the regulatory architecture in GN is shaped by both shared and distinct genetic effects across different disease subtypes. Although most QTLs were shared across at least two GN forms, a fraction of eGenes (5-6%), sGenes (8%), and edQTLs (3-14%) were phenotype-specific, suggesting context-dependent regulatory effects in blood. While 80% of shared eGenes across traits appeared to be driven by the same causal variant by colocalization (PP4 >= 0.8), a measurable subset (2%) showed strong evidence for distinct causal variants (PP3 >= 0.8), implying convergent regulation of the same gene via different regulatory elements depending on disease context. The *RUNX3* example, where different non-colocalized lead variants across FSGS and IgAN map to distinct cis-regulatory elements, illustrates how the same e-gene can be regulated through distinct regulatory architecture depending on the disease context.

Our study also adds resolution by inferring cell-type specificity from bulk transcriptomics. Interaction eQTL mapping using deconvolved cell fractions identified hundreds of cell-type-associated regulatory effects, including signals not detectable in standard bulk eQTL scans, underscoring that disease-relevant regulation can be confined to particular immune cells (e.g., monocytes, T cells, B cells, NK cells). The marked differences in the interaction QTL yield across GN forms (e.g., comparatively few in IgAN despite similar power) suggest that disease context affects gene expression differently across cell types.

Furthermore, our analysis of genotype-by-age, eGFR, and proteinuria effects suggested dynamic genetic regulation of blood gene expression that changes over time. Age-dependent effects at genes such as *RBL2*, and disease severity-related interactions provide a framework for understanding how inherited variation may differentially influence immune pathways across the lifespan and disease stages, potentially explaining the heterogeneity observed clinically in kidney disease progression and related comorbidities.

Despite the unique strengths of our study, several limitations must be considered. First, the sample size of our WGS datasets was insufficient to support discovery of novel disease associations. Nevertheless, we were able to replicate previously reported loci, and we generated a comprehensive set of summary statistics that can be used for subsequent meta-analyses. Second, the use of bulk blood RNA-seq, while practical for large-scale cohort studies, necessitates computational deconvolution of cell-type proportions and can miss more subtle signals from less abundant leukocyte subsets. Third, our age-interaction QTL analysis could be potentially confounded by disease subtype and kidney disease severity, but our key results are robust to both GN type and eGFR adjustments. Lastly, our power to detect cell type interaction QTLs, rare variant associations, and allelic-specific expression was limited, especially for IgAV (smallest GN subtype). These limitations could be addressed in the future by single-cell transcriptomic studies of blood immune cells in larger GN cohorts.

In summary, our cross-GN discovery study revealed thousands of blood QTLs absent from GTEx^21^ and enriched in immune pathways, suggesting that patient cohorts capture disease-activated regulatory programs that are not detectable in healthy tissue resources^58^. We demonstrated that integrating GWAS with disease context-specific QTLs improved biological interpretability of the established loci and prioritized multiple candidate molecular mediators of risk alleles. As the sample sizes and power of GWAS for GN continue to increase, this resource provides critical tools for comprehensive functional annotation and improved interpretability of novel GWAS loci.

## Data Sharing Statement

Individual-level RNA-seq and whole-genome sequencing data are available through dbGaP under accession number phs002480.v5.p4. All GWAS, RVAS, QTL, and ASE summary statistics will be publicly available at the Kiryluk Lab website: https://www.columbiamedicine.org/divisions/kiryluk/resources.php.

## Acknowledgements

We would like to thank study participants of GureGN Study, NEPTUNE Study, Columbia CKD Biobank, and Columbia Genomics Consortium for contributing samples and data for this study. Whole genome sequencing was funded by AstraZeneca. RNA sequencing and related analyses were funded by the NIDDK grant RC2DK116690 (KK, MK). The CureGN cohort was funded by NIDDK grants U24DK100845, U01DK100846, U01DK100876, U01DK100866, and U01DK100867. Patient recruitment for the CureGN study was additionally supported by NephCure. The NEPTUNE cohort was funded by NIH grants U54DK083912 (MK) and U2CTR002818, with additional funding and/or programmatic support provided by the University of Michigan, NephCure, Alport Syndrome Foundation, and the Halpin Foundation. Additional funding included K01DK137031 (LL), 5R01DK105124-08 (KK), 1R21HD104176-01 (RG), 1U01AI152585-01 (RG), 1R01DK134347-01 (RG) and R01DK082753 (AG).

## Disclosures

Generation of WGS data was funded by AstraZeneca. D.S.P and S.P. are employed by AstraZeneca. K.K. reports consulting for Otsuka, Vera, Biogen, Vertex, Travere, Natera, and Novartis. M.K. reports grants and contracts through the University of Michigan outside of this work from Chan Zuckerberg Initiative, Breakthrough T1D, Alport Foundation, National Kidney Foundation, Roche-Genentech, Astra-Zeneca, Moderna, European Union Innovative Medicine Initiative, Boehringer-Ingelheim, Travere Therapeutics, Maze therapeutics, Chinook, RenalytixAI, NovoNordisk, Eli Lilly, Gilead, Regeneron, Bayer, Jansen, Sanofi, Dimerix, Vera Therapeutics. He has received consulting fees through the University of Michigan from Abiologics Inc, Sanofi, Vera Therapeutics, Regeneron, CSL Behring, Flagship Biotech, NovoNordisk, Otsuka, Alexion/AstraZeneca, Variant Bio, Novartis. MK has a patent PCT/EP2014/073413, Biomarkers and methods for progression prediction for chronic kidney disease, licensed. MK also has a Royalty sharing agreement for CKD drug development with AstraZeneca. A.S. has received consulting fees from Boerhinger-Ingelheim, research funding from Dimerix, Boerhinger-Ingelheim, NephCure, and the International Society for Glomerular Diseases. M.H. reports grants and contracts through the University of Michigan outside of this work from Takeda Pharmaceuticals, Hi-Bio (acquired by Biogen) and Calliditas Therapeutics and reports consulting for Novartis. The remaining authors have no conflicts to declare.

## Author Contributions

All authors have read and approved the final version of manuscript. K.K. and M.K. conceived the study, secured funding, provided overall supervision of the project, and made the decision to publish the findings. L.L. performed RNA-seq alignment, transcript and splicing quantification, quality control analyses as well QTL analyses and GWAS colocalizations. C.W. performed WGS bioinformatic processing, variant calling, quality control and ancestry analyses, common and rare variant genetic association scans, and polygenic risk score validations. O.K. performed ASE analyses. F.Z. performed clinical correlation analyses. L.L., D.F. and Q.L. performed double-strand A-to-I RNA editing quantification and editing QTL mapping. D.F. and Y.O. contributed to colocalization analysis. J.Y.Z. contributed to sample and data management. D.F. and F.E. assisted with RNA-seq QC analyses. D.S.P., S.P., D.G. and A.G. facilitated generation and consulted on the analysis of WGS data. All other authors provided analytical and/or clinical feedback through participation in the CureGN Genomics Workgroup.

## Columbia Genomics Consortium

The Columbia Genomics Consortium includes control genomic data from a number of projects and contact investigators: the Columbia University Biobank (Muredach Reilly), the Columbia CKD Biobank (Krzysztof Kiryluk and Ali Gharavi), Genomic Translation for ALS Care (GTAC; Matthew Harms), Pulmonary Fibrosis (Christine Garcia), Atopic disease (Joshua Milner) and Prenatal disease (Ronald Wapner). Only the contact investigator(s) for each project are listed above.

## NEPTUNE Consortium

### NEPTUNE Collaborating Sites

*Atrium Health Levine Children’s Hospital, Charlotte, SC*: Susan Massengill*, Layla Lo^#^

*Cleveland Clinic, Cleveland, OH*: Katherine Dell*, John O’Toole*, John Sedor**, Victoria Grange^#^

*Children’s Hospital, Denver, CO:* Bradley Dixon*, Nathan Rogers^#^

*Children’s Hospital, Los Angeles, CA*: Rachel Lestz*, Natalie Esquivias^#^

*Children’s Mercy Hospital, Kansas City, MO*: Tarak Srivastava*, Kelsey Markus^#^

*Cohen Children’s Hospital, New Hyde Park, NY*: Christine Sethna*, Suzanne Vento^#^

*Columbia University, New York, NY:* Pietro Canetta*

*Duke University Medical Center, Durham, NC:* Opeyemi Olabisi*, Rasheed Gbadegesin**, Kimberly Cicio^#^

*Emory University, Atlanta, GA:* Laurence Greenbaum*, Chia-shi Wang*, Chris Fan^#^

*The Lundquist Institute, Torrance, CA:* Sharon Adler*, Janine LaPage^#^

*John H Stroger Cook County Hospital, Chicago, IL:* Amatur Amarah*

*Johns Hopkins Medicine, Baltimore, MD:* Meredith Atkinson*, Ryan Hutson^#^

*Mayo Clinic, Rochester, MN:* John Lieske, Marie Hogan, Fernando Fervenza

*Medical University of South Carolina, Charleston, SC:* David Selewski*, Cheryl Alston^#^

*Montefiore Medical Center, Bronx, NY:* Kim Reidy*, Michael Ross*, Frederick Kaskel**, Patricia Flynn^#^

*New York University Medical Center, New York, NY:* Laura Malaga-Dieguez*, Olga Zhdanova**, Laura Jane Pehrson^#^, Melanie Miranda^#^

*The Ohio State University College of Medicine, Columbus, OH*: Salem Almaani*, Laci Roberts^#^

*Riley Children’s Hospital of Indiana University, Indianapolis, IN:* Myda Khalid*, Veronica Servin^#^

*Stanford University, Stanford, CA:* Richard Lafayette*, Elizabeth Chen^#^

*Temple University, Philadelphia, PA:* Iris Lee**

*Texas Children’s Hospital at Baylor College of Medicine, Houston, TX*: Shweta Shah*, Thinh Phan^#^

*University Health Network Toronto:* Heather Reich*, Michelle Hladunewich**, Paul Ling^#^, Martin Romano^#^

*University of California at San Diego, San Diego, CA:* Ambarish Athavale*, Caitlin Carter*, Kristin Zeeb^#^

*University of California at San Francisco, San Francisco, CA*: Paul Brakeman*, Daniel Schrader

*University of Colorado Anschutz Medical Campus, Aurora, CO*: James Dylewski* Nathan Rogers^#^

*University of Kansas Medical Center, Kansas City, KS*: Ellen McCarthy*, Catherine Creed^#^

*University of Miami, Miami, FL:* Alessia Fornoni*, Miguel Bandes^#^

*University of Michigan, Ann Arbor, MI:* Matthias Kretzler*, Laura Mariani*, Zubin Modi*, Amanda Williams^#^, Roxy Ni^#^

*University of Minnesota, Minneapolis, MN:* Patrick Nachman*, Michelle Rheault*, Ariel Langenberger^#^, Brady Wallner^#^

*University of North Carolina, Chapel Hill, NC:* Vimal Derebail*, Keisha Gibson*, Anne Froment^#^, Sharia Warren^#^

*University of Pennsylvania, Philadelphia, PA:* Lawrence Holzman*, Kevin Meyers**, Krishna Kallem^#^, Arielle Swenson^#^

*University of Texas San Antonio, San Antonio, TX*: Samin Sharma**

*University of Texas Southwestern, Dallas, TX:* Elizabeth Roehm*, Kamalanathan Sambandam**, Elizabeth Brown**

*University of Washington, Seattle, WA:* Ashley Jefferson*, Sangeeta Hingorani**, Katherine Tuttle^**§^, Linda Manahan ^#^, Emily Pao^#^, Kelli Kuykendall^§^

*Wake Forest University Baptist Health, Winston-Salem, NC:* Jen Jar Lin**

*Washington University in St. Louis, St. Louis, MO*: Brian Stotter*, Joseph Dumayas^#^

### Data Analysis and Coordinating Center

*University of Michigan:* Matthias Kretzler*, Brenda Gillespie**, Laura Mariani**, Zubin Modi**, Eloise Salmon**, Howard Trachtman**, Hailey Desmond, Sean Eddy, Damian Fermin, Wenjun Ju, Maria Larkina, Chrysta Lienczewski, Rebecca Scherr, Jonathan Troost, Amanda Williams, Yan Zhai; *Cleveland Clinic:* Crystal Gadegbeku**, John Sedor**, *Duke University:* Laura Barisoni**; *Harvard University:* Matthew G Sampson**; *Northwestern University:* Abigail Smith**; *University of Pennsylvania:* Lawrence Holzman**, Jarcy Zee**

### Digital Pathology Committee

Carmen Avila-Casado *(University Health Network)*, Serena Bagnasco *(Johns Hopkins University)*, Lihong Bu *(Mayo Clinic)*, Shelley Caltharp *(Emory University)*, Clarissa Cassol *(Arkana)*, Dawit Demeke *(University of Michigan)*, Brenda Gillespie *(University of Michigan)*, Jared Hassler *(Temple University)*, Leal Herlitz *(Cleveland Clinic)*, Stephen Hewitt *(National Cancer Institute)*, Jeff Hodgin *(University of Michigan)*, Danni Holanda *(Arkana)*, Neeraja Kambham *(Stanford University)*, Kevin Lemley, Laura Mariani *(University of Michigan)*, Nidia Messias *(Washington University)*, Alexei Mikhailov *(Wake Forest)*, Vanessa Moreno *(University of North Carolina)*, Behzad Najafian *(University of Washington)*, Matthew Palmer *(University of Pennsylvania)*, Avi Rosenberg *(Johns Hopkins University)*, Virginie Royal *(University of Montreal)*, Miroslav Sekulik *(Columbia University)*, Barry Stokes *(Columbia University)*, David Thomas *(Duke University)*, Ming Wu *(University of New York)*, Michifumi Yamashita *(Cedar Sinai)*, Hong Yin *(Emory University)*, Jarcy Zee *(University of Pennsylvania)*, Yiqin Zuo *(University of Miami)*. Co-Chairs: Laura Barisoni *(Duke University)*, Cynthia Nast *(Cedar Sinai)*.

## CureGN Consortium

The CureGN Consortium members listed below, from within the four Participating Clinical Center networks and Data Coordinating Center, are acknowledged by the authors as Collaborators.

**CureGN PCC Principal Investigators; *CureGN Site Principal Investigators; ^+^CureGN Pathologists, ^#^CureGN Lead Coordinators.

### CureGN Participating Clinical Centers (PCC) through Columbia University

*Columbia University, New York, NY, US*: Gerald Appel, Revekka Babayev, Ibrahim Batal ^+^, Andrew Bomback**, Pietro Canetta, Brenda Chan, Vivette Denise D’Agati ^+^, Samitri Dogra, Hilda Fernandez, Gabriele Gaggero^+^, Ali Gharavi**, William Hines, , Krzysztof Kiryluk**, Satoru Kudose ^+^, Fangming Lin, Victoria Kolupaeva#, Maddalena Marasa, Glen Markowitz ^+^, Mariela Navarro-Torres, Hila Milo Rasouly, Sumit Mohan, Nicola Mongera, Jordan Nestor, Jai Radhakrishnan, Maya Rao, Maya Sabatello, Simone Sanna-Cherchi, Dominick Santoriello^+^, Miroslav Sekulic ^+^, Michael Barry Stokes^+^, Natalie Uy, Natalie Vena, Benjamin Wooden

*University of Warsaw, Warszawa, Poland:* Bartosz Foroncewicz, Natalia Wiewiórska-Krata, Barbara Moszczuk, Krzysztof Mucha*, Agnieszka Perkowska-Ptasińska, Elżbieta Ryszkowska

*IRCCS Giannina Gaslini, Genoa, Italy*: Francesca Lugani, Valerio Vellone^+^

### CureGN Participating Clinical Centers (PCC) through the Pediatric Nephrology Research Consortium

*Children’s Hospital of New Orleans/ LSU Health, New Orleans, LA, USA*: Diego Aviles*

*Children’s Mercy Hospital, Kansas City, MO, USA*: Tarak Srivastava*, Alexander Katz^+^

*Children’s National Medical Center, Washington DC, USA*: Sun-Young Ahn*

*Cincinnati Children’s Hospital Cincinnati, OH, USA*: Prasad Devarajan, Elif Erkan*, Hillarey Stone

*Connecticut Children’s Medical Center, Hartford, CT, USA*: Sherene Mason*

*East Carolina University Brody School of Medicine, Greenville, NC, USA*: Liliana Gomez-Mendez*

*Emory University, Atlanta, GA, USA*: Larry Greenbaum**, Chia-shi Wang, Hong (Julie) Yin^+^

*Helen DeVos Children’s Hospital, Grand Rapids, MI, USA*: Shruthi Mohan*

*Levine Children’s Hospital/Atrium Health, Charlotte, NC, USA*: Donald Weaver*

*Lurie Children’s Hospital, Chicago IL, USA*: Jill Krissberg*, Jerome Lane

*Medical College of Wisconsin, Milwaukee, WI, USA*: Cindy Pan, Ellen Cody*

*Nationwide Children’s Hospital, Columbus, OH, USA*: Dawson Carmean^#^, Mahmoud Kallash*, John Mahan**, Samantha Sharpe^#^, William Smoyer**, Laura Biederman^+^, Desirée Jones ^#^

*Oregon Health and Science University, Portland, OR, USA*: Chloe Douglas*, Sandra Iragorri

*Riley Children’s Hospital, Indianapolis, IN, USA*: Myda Khalid**

*Cardinal Glennon Children’s Medical Center/ St. Louis University, St. Louis, MO, USA*: Craig Belsha*

*Texas Children’s Hospital, Houston, TX, USA*: Elizabeth Onugha*, Michael Braun, AC Gomez

*Children’s of Alabama, University of Alabama, Birmingham, AL, USA*: Daniel Feig*

*University of Colorado Children’s Hospital, Colorado, Aurora, CO, USA*: Melisha Hannah*

*University of Kentucky, Lexington, KY, USA*: Aftab Chishti*

*University of Louisville, Louisville, KY, USA*: Jon Klein**

*Holtz Medical Center, University of Miami, Miami, FL, USA*: Chryso Katsoufis, Wacharee Seeherunvong*

*University of Minnesota Children’s Hospital, Minneapolis, MN, USA*: Michelle Rheault**

*University of New Mexico Health Sciences Center, Albuquerque, NM, USA*: Craig Wong*

*University of Oklahoma Health Sciences Center, Oklahoma City, OK, USA*: Qassim Abid*

*University of Virginia, Charlottesville, VA, USA*: John Barcia*, Agnes Swiatecka-Urban

*University of Wisconsin, Madison, WI, USA*: Sharon Bartosh*

*Washington University in St. Louis, St. Louis, MO, USA*: Kaye Brathwaite*, Joseph Gaut ^+^

CureGN Participating Clinical Centers (PCC) through the University of North Carolina:

*Hôpital Maisonneuve-Rosemont, Montreal, Canada*: Louis-Philippe Laurin*, Virginie Royal^+^, Mathieu Latour^+^, Natlie (Natacha) Patey ^+^

*Medical University of South Carolina, Charleston, SC, USA*: Anand Achanti, Milos Budisavljevic*, Vishwajeeth Pasham^+^

*Northwestern University, Chicago, IL, USA*: Cybele Ghossein, Yonatan Peleg*

*Ohio State University, Columbus, OH, USA*: Isabelle Ayoub*, Samir Parikh, Brad Rovin, Anjali Satoskar^+^

*University of Chicago, Chicago, IL, USA*: Anthony Chang^+^

*University of Alabama at Birmingham, Birmingham, AL, USA*: Huma Fatima^+^, Jan Novak, Matthew Renfrow, Dana Rizk*

*University of North Carolina Kidney Center, Chapel Hill, NC, USA*: Dhruti Chen, Vimal Derebail**, Ronald Falk**, Keisha Gibson, Dorey Glenn, Susan Hogan, Koyal Jain, J. Charles Jennette^+^, Vanessa Moreno^+^, Amy Mottl, Caroline Poulton^#^, Monica Reynolds, Manish Kanti Saha, Nicole E. Wyatt

*Vanderbilt University, Nashville, TN, USA*: Agnes Fogo^+^, Yasar Caliskan*

*Virginia Commonwealth University, Richmond, VA, USA*: Jason Kidd*, Selvaraj Muthusamy^+^

### CureGN Participating Clinical Centers (PCC) through the University of Pennsylvania

*Children’s Hospital of Philadelphia, Philadelphia, PA, USA*: Rebecca Scobell*, Michelle Denburg, Amy Kogon, Kevin Meyers, Madhura Pradhan

*Cleveland Clinic, Cleveland, OH, CA*: Raed Bou Matar*, John O’Toole, John Sedor

Cohen Children’s Medical Center, New Hyde Park, NY, USA: Christine Sethna**, Suzanne Vento^#^

*Johns Hopkins University, Baltimore, MD, USA*: Mohamed Atta, Serena Bagnasco^+^, Alicia Neu, John Sperati*

*Lundquist Institute at Harbor-UCLA Medical Center, Torrance, CA, USA*: Sharon Adler*, Tiane Dai, Ram Dukkipati

*Montefiore Medical Center, The Bronx, New York, NY, USA*: Frederick Kaskel, Kimberly Reidy*

*New York University, New York, NY, USA*: Laura Malaga-Dieguez*

*Spokane Providence Medical Center, Spokane, WA, USA*: Katherine Tuttle*

*Stanford University, Palo Alto, CA, USA*: Richard Lafayette*, Kamal Fahmeedah, Elizabeth Talley

*Sunnybrook Health Sciences Centre, Toronto, Canada*: Michelle Hladunewich*

*The Hospital for Sick Children, Toronto, Canada*: Rulan Parekh*

*University Health Network, Toronto, Canada*: Carmen Avila-Casado^+^, Daniel Cattran*, Reich Heather, Meherzad Kutky

*University of Miami, Miami, FL, USA*: Yelena Drexler*, Alessia Fornoni

*University of Michigan, Ann Arbor, MI, USA*: Jeffrey Hodgin^+^, Andrea Oliverio*

*University of Pennsylvania, Philadelphia, PA, USA*: Jon Hogan, Lawrence Holzman**, Matthew Palmer ^+^, Gaia Coppock

*University of Pittsburgh School of Medicine, Pittsburgh, PA, USA*: Michael Mortiz, Juhi Kumar*

*University of Washington, Seattle, WA, USA*: Charles Alpers^+^, J. Ashley Jefferson*

*UT Southwestern, Dallas, TX, USA*: Kamal Sambandam, Bethany Roehm*

*University of Minnesota*: Patrick Nachman*

### Data Coordinating Center (DCC)

*Cedar Sinai Medical Center, Los Angeles, CA, USA*: Cynthia Nast^+^, Jean Hou^+^

*Duke University, Durham, NC, USA*: Laura Barisoni

*Cleveland Clinic, Cleveland, OH, USA:* Crystal Gadegbeku**

*Northwestern University, Chicago, IL, USA:* Abigail Smith**

*University of Michigan, Ann Arbor, MI, USA*: Brenda Gillespie, Bruce Robinson, Bethany Klunder,

Matthias Kretzler, Zubin Modi, Laura Mariani**

### Steering Committee Chair

Lisa M. Guay-Woodford, Children’s Hospital of Pennsylvania, Philadelphia, PA, USA

## Methods

### Study cohorts

The genetic cohorts were comprised of 4,307 cases of primary GN defined by a kidney biopsy diagnosis of IgAN, IgAV, MN, MCD, or FSGS and 3,845 ancestry-matched controls without evidence of kidney disease. The cases included 2,007 participants enrolled in the CureGN study^18^, 358 participants enrolled in the NEPTUNE study^59^, 1,942 participants enrolled in the Columbia University CKD Biobank. The controls drawn from the pool of anonymized non-kidney disease participants of other studies sequenced by the Columbia Genomics Consortium and matched based on genetic ancestry to the cases. The demographic and clinical characteristics of the included participants are summarized in **Table 1**. All participants provided consent for genetic studies, and our protocol was approved by the Columbia IRB (protocol #AAAC7385).

**Table 1.** Demographic and clinical characteristics of patients in the study cohort.

|  | **FSGS** | **MCD** | **MN** | **IgAN** | **IgAV** |
| --- | --- | --- | --- | --- | --- |
| **WGS** |  |  |  |  |  |
| No. of patients* | 508 | 606 | 962 | 1,496 | 377 |
| Age, median (Q1-Q3) | 32 (15-49) | 20 (7-44) | 52 (37-64) | 30 (18-43) | 10 (7-16) |
| Male (%) | 266 (53%) | 323 (53%) | 600 (63%) | 893 (60%) | 205 (55%) |
| eGFR, median (Q1-Q3) [mL/min/1.73m<sup>2</sup>] | 71 (43-103) | 102 (73-123) | 90 (62-109) | 72 (43-99) | 101 (73-121) |
| UPCR, median (Q1-Q3) [g/g] | 4.0 (1.9-8.0) | 5.2 (1.1-9.6) | 5.6 (3.1-8.5) | 1.5 (0.8-3.0) | 1.3 (0.5-4.0) |
| **WGS + RNA-seq** |  |  |  |  |  |
| No. of patients | 447 | 408 | 441 | 403 | 123 |
| Age, median (Q1-Q3) | 32 (15-49) | 14 (5-37) | 51 (37-62) | 31 (16-44) | 14 (8-26) |
| Male (%) | 242 (54%) | 218 | 272 (61%) | 244 (60%) | 73 (59%) |
| eGFR, median (Q1-Q3) [mL/min/1.73m<sup>2</sup>] | 71 (42-102) | 105 (78-124) | 89 (64-110) | 69 (43-98) | 92 (65-113) |
| UPCR, median (Q1-Q3) [g/g] | 4.0 (2.1-8.0) | 4.6 (0.7-9.2) | 5.6 (2.9-8.5) | 1.4 (0.7-3.0) | 1.8 (0.7-5.0) |

\* Only newly sequenced individuals are summarized (i.e., the table includes WGS samples from CureGN and the Columbia University CKD Biobank but excludes samples from NEPTUNE).

### Whole genome sequencing and variant calling

We conducted whole genome sequencing using 150 bp paired-end reads on Illumina NovaSeq 6000 instruments, targeting a read depth of 27x or greater. The short germline variant discovery process was performed with the Illumina DRAGEN Bio-IT Platform, comprising sequencing mapping that aligns reads to the Genome Reference Consortium Human Genome Build 37 (GRCh37), and variant calling for individual sample. Variant genotypes were aggregated across all samples employing the joint genotyping tool available in the Genome Analysis Toolkit (GATK) v4.3.0.0^60^. To ensure data quality, we applied variant site filtration to the joint call set using GATK’s variant quality score recalibration (VQSR) model that integrates multiple variant quality metrics, including Fisher strand (FS), mapping quality (MQ), mapping quality rank-sum test (MQRankSum), quality by depth (QD), read position rank-sum test (ReadPosRankSum), and strand odds ratio (SOR)^61^. The cohort-wide low-quality variants were filtered out with VQSR truth sensitivity levels of 99.8% for single nucleotide variants (SNVs) and 99.0% for insertions/deletions (indels). Additionally, we filtered out low-quality genotypes with the following criteria: depth (DP) < 10, genotype quality (GQ) < 20, or allelic balance < 0.2 (or > 0.8) for heterozygotes. Samples were excluded if they had low depth coverage (< 27X), exhibited discordance between self-reported sex and genetic sex (inferred from sex chromosome ploidy calls), had a high contamination rate (FREEMIX score > 3%, as estimated by VerifyBamID^62^), or had low sequence coverage (missing call rate > 20%). To make the genome build consistent with the CureGN RNA sequencing data, the WGS genotype data was mapped to GRCh38 using liftOver^63,64^. The lifted mapping to GRCh38 could improve the downstream genetic analysis when incorporating the most recent reference population datasets and the enhanced annotations of genes and regulatory elements^65^.

### Genetic ancestry and relatedness

Principal component analysis (PCA) was used to estimate the genetic ancestry from a merged study dataset with the WGS reference data from 1000 Genomes Project (1000G)^66^. We conducted PCA on the merged dataset and extracted the first 16 significant principal components (PCs) using FlashPCA2^67^. We employed Louvain clustering on the 16 PCs to broadly classify individuals into three distinct ancestry clusters, as visualized in **Figure S1**. Genetic association testing was performed within each of the three ancestry strata separately, PCA was repeated within each ancestry clusters and five significant PCs determined by Tracy-Widom test were used as covariates in genetic association testing. We deliberately retained low granularity in cluster definitions to maintain a reasonable number of samples per cluster for the downstream association studies. In addition to population structure, we performed relatedness analysis using the KING kinship inference algorithm^68^. We excluded 444 related samples (17/427 cases/controls) to ensure that no pair of individuals exhibited a second-degree or closer relationship (kinship coefficient >= 0.0884). Linear mixed models and stacked ridge regression are widely used to account for the sample relatedness and population structure^69–74^. Nevertheless, these methods tend to be conservative, particularly when the study sample size is limited or when case-control imbalance exist^72,73^. To mitigate the deflation in test statistics in GWAS, we employed a linear model after excluding the related samples. For comparison, we also utilized the linear mixed model on the entire dataset encompassing both unrelated and related samples.

### Genome-wide association studies (GWAS)

For each of the three ancestry clusters, we performed single-variant association tests for the high-quality common variants. In particular, variants with call rate < 0.95, ancestry MAF < 1%, or Hardy-Weinberg equilibrium P<1.0×10^−6^ in ancestry controls were filtered out. We applied the logistic regression model implemented in PLINK2^75^ for ancestry-specific GWAS on the unrelated samples controlling for sex and five PCs derived from each ancestry group separately to account for population structure. In comparison, we also utilized REGENIE^73^ with the related samples included in the analysis. REGENIE is a robust regression-based approach that accounts for sample relatedness, population structure, and case-control imbalance in genetic association analyses. A trans-ancestry analysis was performed to aggregate the ancestry-specific summary statistics using the inverse-variance weighted fixed-effect meta-analysis from METAL^76^.

### Gene-based and region-based rare variant association tests

We conducted an ancestry-specific gene-based association test on the rare protein-coding variants predicted to be damaging to protein function. Multiple types of variant set aggregation tests in REGENIE, including Burden^77^, SKAT-O^78^, and ACAT^79^, were employed on the full dataset including related samples. An omnibus test integrating p-values from Burden, SKAT-O and ACAT tests was also applied. The analysis was adjusted for sex and five PCs. We also employed Fisher’s exact test (FET) as an alternative approach to variant set testing, known for its robustness in rare variant tests but without accounting for confounders^80^. We applied different MAF filters (<0.01, 0.005, and 0.001), based on the ancestry-specific MAF calculated in the studied cohort and minimal population-specific MAFs from gnomAD v3.1.2^81^. We focused on the predicted loss-of-function (pLoF) and deleterious missense (Dmis) variants. First, variant molecular consequences were annotated using the Ensembl variant effect predictor (VEP)^82^. Further, high-confidence pLoF variants were identified by LOFTEE^83^. Dmis variants were determined based on three broadly used variant pathogenicity predictors: PolyPhen-2^84^, REVEL^85^, and AlphaMissense^86^. Specifically, a missense variant is predicted as deleterious if it meets PolyPhen-2 HumDiv score > 0.953, REVEL score > 0.5, and AlphaMissense pathogenicity score >0.564. We considered pLoF combined with Dmis variants in gene-based testing. Another type of gene-centric test is the collapsing of noncoding rare variants in gene regulatory regions^87^. The regulatory elements for protein-coding genes were mapped based on the activity-by-contact (ABC) model^88,89^. The ABC model integrates enhancer activity and enhancer-gene contact frequency measured from cell-type specific epigenomic data. We merged the ABC predictions across 131 cell types and aggregated rare variants within the union of regulatory elements for each protein-coding gene. Only the putative functional variants with CADD (v1.6)^90^ phred score > 15 were included in burden tests. Statistical significance for gene-centric rare variant aggregation tests was determined using Bonferroni correction for the number of tested genes (∼19,000). Associations with p-value < 2.63×10^−6^ were considered exome-wide significant.

Additionally, we performed rare variant aggregation tests at the gene-set level by collapsing rare deleterious coding variants across predefined biological gene sets. Gene sets were obtained from the Molecular Signatures Database (MSigDB)^91^, including gene ontology and human phenotype ontology categories, curated biological pathways, and immunologic signature gene sets, comprising a total of 28,718 gene sets. For each gene set, rare deleterious variants were aggregated across all protein-coding genes included in the corresponding set. Rare deleterious variants were defined using the same criteria as those applied in the gene-based association tests. MSigDB gene sets are highly overlapping and therefore statistically correlated. We estimated the effective number of independent tests using an eigenvalue-based approach^92^. Briefly, eigen decomposition was performed on the gene-set correlation matrix. The effective number of gene sets was then defined as the minimum number of principal components required to explain 95% of the cumulative eigenvalue variance. Statistical significance for gene-set association analyses was determined using Bonferroni correction based on the effective number of independent tests. The effective number of gene sets was estimated to be 8,392, corresponding to a significance threshold of 5.96×10^−6^.

We also applied the fixed-size sliding window scanning test genome-wide^93–96^ to aggregate rare variants with MAF < 0.01 (< 0.005, < 0.001) and CADD phred score > 15 in each sliding window. Specifically, we applied the variant set tests to 2kb, 5kb, 10kb, 20kb, and 50kb sliding windows, with half of the window overlapping with adjacent windows on each side. The windows start from the beginning of chromosomes and cover the whole genome, providing comprehensive screening for all the possible functional regions. Although the sliding window method provides extensive coverage, the boundaries of testing regions are arbitrary, which may not fully reflect the underlying biological functions of variant sets. As an alternative, we performed the region-based tests by aggregating rare functional variants within the putative candidate cis-regulatory elements (cCREs) using the SCREEN portal V3 (https://screen.encodeproject.org/), which compiled cCRE annotations across 1,518 cell-type-specific epigenetic signatures from ENCODE^38^. The rare functional variants within each cCRE region were collapsed to derive the region-based tests.

### Polygenic risk scores

We calculated the existing polygenic risk scores (PRS) for glomerular diseases and evaluated their predictive performance in the CureGN study. The published models reported in the latest GWAS studies for IgAN^5^, MN^9^, and SSNS^7^ were calculated and tested using our WGS dataset. Unlike genome-wide PRS for IgAN and MN, genome-wide PRS for SSNS has not been previously optimized, thus we optimized model parameters using 300 MCD cases randomly selected as the training set from the Columbia University CKD Biobank and the NEPTUNE projects along with 600 ancestry-matched control samples. Using LDpred^97^ and P+T^98^ methods, multiple PRS models were calculated based on the published trans-ancestry GWAS summary statistics^7^. The model with the highest Nagelkerke’s pseudo-R² was selected for downstream analyses in non-overlapping testing datasets. We also applied previously published 30-SNP IgAN^5^ and the 6-SNP MN^9^ genetic risk scores to compare the performance with genome-wide predictors. Given diverse ancestral background of our cohort, we applied the ancestry calibration method as proposed by us previously^98^. We evaluated predictive performance by applying the PRS to discriminate each glomerular disease subtype from healthy controls as well as from disease controls (i.e., all other unrelated GN types). Predictive performance was assessed using receiver operating characteristic (ROC) curves, with the area under the curve (AUC) serving as a summary measure of discrimination.

Clinical correlation analyses were performed in CureGN participants with WGS data (N=2,010). Three best-performing disease-specific scores were evaluated, each within its corresponding glomerular disease subgroup: the genome-wide PRS for IgAN tested in IgAN cases (N=439) and in the combined IgAN / IgAV group (N=567), the 6-SNP MN GRS in MN cases (N=485), and the genome-wide PRS for SSNS in MCD cases (N=445). For each subgroup, we tested the association between the score and clinical, laboratory and histologic parameters: age at diagnosis, infections at disease onset, estimated glomerular filtration rate (eGFR), urinary protein-to-creatinine ratio (uPCR) and hematuria at the time of biopsy; treatment-response phenotypes (infrequent relapsing, frequent relapsing, steroid resistant, and multidrug resistant), and kidney-biopsy findings, including PLA2R positivity and immunofluorescence C3 intensity in MN, and IgA immunofluorescence intensity and MEST-C scores in IgAN. All models were adjusted for sex, age and the first five genetic principal components. Per standard-deviation increment of the score, effect sizes are reported as odds ratios (OR) with 95% confidence intervals for binary outcomes and as regression coefficients (β) with 95% confidence intervals for continuous and count outcomes, with two-sided P < 0.05 considered nominally significant. We additionally evaluated kidney disease progression using Cox proportional-hazards models. The lifetime risk of end-stage kidney disease (ESKD) was modeled with age as the time scale, adjusting for sex, the first five principal components, and *APOL1* risk genotype. The composite outcome of time from biopsy to ESKD or a 40% decline in eGFR was assessed with adjustement for age at biopsy, sex, the first five ancestry PCs, *APOL1* risk genotype, and eGFR and uPCR at the time of biopsy. Hazard ratios (HR) with 95% confidence intervals were reported per standard-deviation increment of the score. All analyses were performed in R (version 4.6.0).

### Whole blood RNA-sequencing and transcript quantification

Peripheral whole blood was obtained from CureGN participants using PAXgene RNA collection tubes. Samples were maintained at ambient temperature for two hours immediately following collection, then stored frozen at −80C. The RNA extractions were performed using standard protocols. Briefly, the samples were centrifuged to separate cellular material, and the resulting fractions were subjected to lysis, enzymatic digestion, and homogenization with the QuickGene RNA Blood Cell Kit S (RB-S). RNA isolation was carried out on the QuickGene 810 Nucleic Acid Isolation System, followed by additional purification using the Zymo Research RNA Clean & Concentrator-5 kit (R1014). For transcriptome profiling, total RNA samples demonstrating high integrity (RIN > 8; input range 200 ng-1 µg) underwent poly(A) selection to enrich in messenger RNA. Sequencing libraries were constructed with the Illumina TruSeq RNA Library Preparation Kit and sequenced at the Columbia University Genome Center on Illumina HiSeq 2500 and HiSeq 4000 platforms. This approach generated 100-base paired-end reads, with a median sequencing depth of 109.7 million reads per sample. Primary data processing included base calling with Illumina’s Real-Time Analysis (RTA) software and conversion of BCL files to FASTQ format using bcl2fastq2 (v2.17), incorporating adapter trimming. Reads were aligned to the human reference genome (hg38) using STAR^99^. Quality control procedures followed established GTEx consortium guidelines^21^: samples were excluded if they contained fewer than 10 million mapped reads, exhibited a mapping rate below 20%, showed intergenic mapping above 30%, had a base mismatch rate exceeding 1%, or demonstrated rRNA mapping greater than 30%. Additional outlier detection was performed using correlation-based expression metrics and sex concordance checks were performed against records as well as against WGS. Gene-level quantification was performed with RNA-SeQC^100^, generating both raw read counts and transcripts per million (TPM) values. Transcripts were retained for downstream analyses if they achieved TPM > 0.1 and at least 6 read counts in a minimum of 20% of samples. After all quality control steps, the final dataset comprised of 1,822 individuals, including 447 FSGS, 441 MN, 408 MCD, 403 IgAN and 123 IgAV cases.

### Expressing QTL (eQTL) mapping

For eQTL mapping, gene counts were normalized across samples using Trimmed Mean of M-values (TMM) method^101^, and the expression values for each gene were inverse-normal transformed. The cis-eQTLs were identified using linear regression as implemented in tensorQTL^102^, adjusting for age, sex, top 5 ancestry PCs and PEER factors derived from gene expression matrix. We adjusted 15 PEER factors in IgAV cohort and 60 PEER factors in the FSGS, IgAN, MCD, and MN cohorts. We further used CIBERSOFTx^103^ to deconvolve immune cell type abundance from bulk blood RNA-seq of 1,822 CureGN participants. We optimized this pipeline for 5 different major immune cell types, including CD4 and CD8 T cells, NK cells, monocytes, and B cells, using published reference panels generated by scRNA-seq of 25,000 blood cells from 45 healthy donors^25^. The deconvolution was conducted using the default parameters. We adjusted eQTL analyses for the estimated cell fractions (CF) to account for variability in cell composition across individuals. A false discovery rate (FDR) threshold ≤ 0.05 was applied to identify genes with at least one significant cis-eQTL (eGenes). To identify the list of all significant variant-gene pairs associated with cis-eGenes, a genome-wide empirical P-value threshold was defined as the empirical P-value of the gene closest to the FDR 0.05 threshold, as descripted previously^104^. For each gene, variants with a nominal P-value below the gene-level threshold were considered significant and included in the final list of variant-gene pairs. We performed the eQTL mapping for each GN type separately, followed by all combined analysis with additional adjustment for GN type.

### Splicing quantification and splicing QTL (sQTL) mapping

Splicing quantification was based on the intron excision rates computed by LeafCutter^26^. Introns with few counts or with insufficient variability across samples were removed as described previously^21^. The filtered counts were normalized using the LeafCutter and the resulting splicing quantification files were used for sQTL mapping. Mapping of cis-sQTLs was conducted separately within each GN type using tensorQTL^102^ controlling for age, sex, top 5 ancestry PCs, and 15 splicing PEER factors as described in^21^. We also performed a joint cross-phenotype sQTL mapping, including GN type as another covariate. Consistent with the eQTL analysis framework, we also performed sQTL mapping with further adjustment for estimated cell-type fractions. To identify cis-sGenes, FDR was calculated in the same manner as for cis-eQTL, and an sGene-level nominal P-value threshold was determined for all significant variant-intron excision pairs.

### RNA-editing quantification and editing QTL mapping

To quantify editing levels, we used a list of 2,802,572 human RNA editing reference sites, incorporating established sites from the RADAR database^105^, tissue-specific sites identified in GTEx V6p17^106^, and recently reported hyper-editing sites^107^. At each of the reference editing site, we computed the ratio of G reads divided by the sum of A and G reads, based on the RNA-seq samples with matching genotyping data from 1,822 CureGN participants. We only keep the editing sites covered by more than 20 reads in at least 20% of samples and exhibiting non-zero variability in editing levels across samples^27^. We obtained 21,547-25,454 editing sites across GN forms. For edQTL mapping, we considered SNPs with MAF > 0.05 within ±100 kb of editing sites. Raw editing-level measurements were logit-transformed and then inverse normal transformed across individuals within each phenotype^27^. We used linear regression model implemented in tensorQTL^102^, controlling for age, sex, top 5 genetic PCs and top 15 principal components derived from the editing level matrix^22^. An FDR threshold ≤ 0.05 was applied to identify editing sites with at least one significant edQTL (edGenes). To identify the list of all significant variant-editing site pairs associated with edGenes, a genome-wide empirical P-value threshold was defined as the empirical P-value of the site closest to the 0.05 FDR threshold as descripted previously^104^. For each editing site, variants with a nominal P-value below the site-level threshold were considered significant and included in the final list of variant-gene pairs. We performed the edQTL mapping for each GN type separately, followed by all combined analysis with additional adjustment for the disease type.

### Allele-specific Expression (ASE)

We performed ASE analysis in two stages. First, allelic counts per gene per sample were generated. Second, we aggregated these counts to obtain population-level ASE statistics and identify significant ASE signals. For the first stage, we used the phASER software^108^. For the population-level aggregation, we adapted the framework implemented in phASER-POP^109^. Prior to ASE quantification, genotype data were phased using SHAPEIT5^110^. Variants with MAF ≥ 0.1% were phased first in 20 cM chunks to establish a reliable haplotype backbone. Rare variants (MAF<0.1%) were subsequently phased in smaller chunks of 4 cM using phased common variants as a haplotype scaffold^110^. The resulting phased genotypes were then used for haplotype-resolved ASE quantification with phASER^108^. For each sample, read-backed phasing was used to link nearby heterozygous variants covered by the same RNA sequencing reads into haplotype blocks, which were subsequently anchored to the genome-wide phase. In contrast to the population-based phasing, rare variants can provide useful information here because they may be linked to nearby heterozygous sites through read-level evidence. Gene-level allelic read counts were then generated for each sample using the GeneAE module of phASER^108^. Because allelic expression can only be quantified from reads overlapping heterozygous variants, the detectable allelic gene coverage in a given sample depends on the number of heterozygous sites present within that gene. Consequently, genes with few heterozygous variants tend to have reduced allelic read coverage and lower power for ASE detection, a known limitation of ASE analyses based on short-read RNA-seq^36^.

To validate eQTL results, we computed population-level ASE statistics for the top variant-gene pairs identified in CureGN using phASER-POP^109^. For each gene, we selected a single top variant corresponding to the most significant association signal, resulting in one variant per gene mapping. The first step of the phASER-POP workflow aggregates per-sample allelic expression estimates into a matrix of phased allelic counts across all samples. In the second step, ASE is evaluated for each variant-gene pair by comparing |log aFC| (absolute value of the log allelic fold change) distributions between heterozygous and homozygous samples. This value is a measure of allelic imbalance in each sample, and the usual expectation is that when the allele-specific regulation is present, the absolute log aFC values would be larger in heterozygous samples compared to the homozygous ones. A Wilcoxon two-sided rank-sum test is performed by phASER-POP to compare the absolute log aFC values between the heterozygous and homozygous samples, but since we are interested only in one side of this comparison (under the hypothesis that |log aFC| values are *larger* in heterozygous samples), we convert:

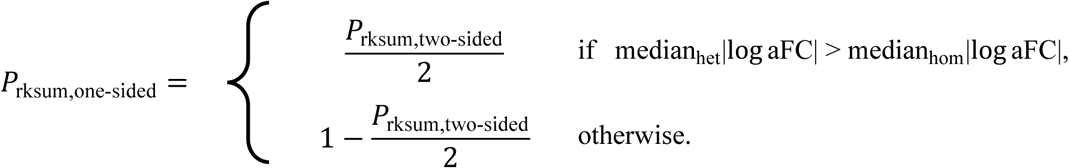

The rank-sum statistic can be biased due to differences in detectable allelic expression between heterozygous and homozygous samples. Because allelic reads are only measurable at heterozygous sites, genes with more heterozygous variants tend to have higher observed allelic counts. Samples heterozygous at the test variant are also more likely to carry additional heterozygous variants within the gene due to linkage disequilibrium, increasing measured allelic expression in heterozygous samples relative to homozygous samples. Conversely, lower coverage in homozygous samples inflates the dispersion of |log aFC| values, which can bias the rank-sum statistic in the opposite direction and render the rank-sum-based one-sided p-values overly conservative. Consistent with this bias, up to ∼40% of signals significant under the two-sided test (after multiple-testing correction) exhibited larger |log aFC| values in homozygous samples, a pattern without clear biological interpretation. To overcome these limitations, we implemented a complementary ASE significance measure based solely on heterozygous samples. In this approach, the sign of the log aFC phased according to the test variant is used to determine the direction of allelic imbalance. Under the null hypothesis of no cis-regulatory effect, positive and negative signs occur with equal probability, yielding a binomial distribution with success probability 0.5. Thus, we define the corresponding p-value using binomial statistics with parameters n, the number of all heterozygous samples, and p=0.5, based on the observed number of positive signs across heterozygous samples. Because this approach depends only on the direction of imbalance within each sample, it is robust to differences in total allelic coverage and independent of sample-level covariates. The contrast between rank-sum-based and binomial-based statistics is illustrated in **Figure S12**: extreme dispersion among homozygous samples can lead to misleading rank-sum results, while the binomial test consistently detects ASE effects in heterozygous samples. In the Results section, we refer to the rank-sum-based approach as the *magnitude-based* approach, as it captures differences in the magnitude of allelic imbalance, and the binomial-based approach as the *direction-based* approach, as it quantifies the consistency of imbalance direction across samples.

### Interaction QTL mapping

We used CIBERSOFTx^103^ deconvolved cell fractions of 5 major cell types as described above for interaction eQTL mapping (ieQTLs) to investigate eQTL effects specific to different cell types in each GN type. The ieQTL mapping for each cell type was performed using tensorQTL testing variants within ±1Mb of the TSS of each gene. Given modest sample size for interaction testing, we restricted ieQTL mapping to variants with MAF ≥ 0.05 to avoid potential regression outlier effects. Similarly, we tested ieQTLs by incorporating eGFR and proteinuria as interaction terms. A gene with at least one significant ieQTL was defined as an ieGene. To identify ieGenes, the top interaction p-value for each gene was corrected for multiple testing using eigenMT. Significance across genes was computed by adjusting the eigenMT-corrected p-values using Benjamini-Hochberg and applying FDR threshold of 0.05.

### Gene set enrichment analyses

Gene set enrichment analyses were performed using ToppFun^111^, a functional enrichment module of the ToppGene Suite. The prioritized genes were submitted as input gene lists and analyzed for enrichment across curated annotation categories, including Gene Ontology, mouse phenotype, pathway databases, and other functional annotations available in ToppFun. Enrichment was performed using the default hypergeometric-based statistical framework with the transcriptome-wide gene set as the background. Resulting P values were corrected for multiple hypothesis testing using the Benjamini-Hochberg false discovery rate (FDR) method, and enriched terms with an FDR below 0.05 were considered statistically significant.

### Colocalization between traits

Colocalization analyses were performed using COLOC^112^ under default parameters. For each trait pair, we estimated the posterior probability of a shared causal variant, and a “colocalization” was defined based on the posterior probability for the hypothesis 4 of shared causal variant (PP4) greater than 80%^113^. We considered traits to have distinct causal variants when the posterior probability for hypothesis 3 (PP3: association with trait 1 and trait 2, two independent SNPs) greater than 80%. To test the shared and distinct genetic regulation between traits, we performed colocalization at each of shared eGene to each pair of traits. We further conducted colocalization analyses at each GWAS locus associated with glomerular disorders^2–7,9–13^ with corresponding identified QTL signals, including eQTL, sQTL and edQTL, to prioritize candidate causal molecular mediators and underlying regulatory mechanisms.

## Notes

### Competing Interest Statement

The authors have declared no competing interest.

### Author Declarations

Columbia University IRB approved this work.

### Summary of Updates

We updated our analysis of GWAS and QTL integration and added Figure 6 as an additional main figure.

