## Supplementary Information for "Atlas of glomerular disease-specific genetic effects on blood transcriptome"

*Liu et al.*

**Supplementary Results**

**Blood immune cell type-specific genetic effects**

Because blood QTL mapping with bulk RNA-seq may potentially obscure the cellular specificity of genetic regulatory mechanisms, we next performed interaction eQTL (ieQTL) mapping across major blood cell types using computational estimates of cell type proportions to identify cell type–specific eQTL signals^1^. The bulk blood gene expression data were deconvoluted into five major immune cell types, including B cells, CD4 T cells, CD8 T cells, Monocytes and NK cells^2,3^. The ieQTL mapping was conducted to test an interaction between genotype and each cell type fraction using a linear regression model, controlling for age, sex, genetic ancestry, and latent factors in the expression data.

Across cell types and GN types, we detected 530 protein coding and lincRNA genes with an ieQTL (ieGenes) at 5% FDR per GN-cell type combination (**Figure S7 and Table S8-S12**). About 90% of ieQTLs corresponded to genes with at least one cis-eQTL across GN types mapped in bulk blood tissue, whereas 10% of these ieQTLs were not detected by standard eQTL analysis. The MCD cohort exhibited the highest number of ieQTLs (N=289), followed by FSGS (N=242) and MN (N=147). Notably, in contrast to standard eQTL signals, a large proportion of identified cell type-specific ieQTLs were GN context-specific (**Figure S7**).

**Allele-specific expression**

Allele-specific expression (ASE) analysis provides an orthogonal complement to cis-eQTL mapping by leveraging reads overlapping heterozygous sites to quantify allelic imbalance within the same individual and can be used to validate cis-eQTL signals^4,5^. Because the two allelic measurements are obtained from the same sample and processed together, ASE is inherently robust to many sample-level factors that affect total expression similarly for both alleles, including environmental and technical effects. We used ASE primarily to validate cis-eQTL signals across the GN phenotypes. For each variant, gene, and sample, we quantified allelic imbalance as the phased log allelic fold change (aFC) between the two alleles using phASER^6^ (**Methods**). We used 5% FDR to define significant allele-specific expression QTLs (aseQTLs) across the transcriptome.

Comparison of significant genes between ASE and cis-eQTL analyses showed that most aseQTL signals overlap with cis-eQTL signals (**Figure S9a**). Overall, >92% of RankSum-based (referred to as magnitude-based) aseQTLs were also detected as cis-eQTLs, and >81% of Binomial-based (referred to as direction-based) aseQTLs were cis-eQTLs; in non-IgAV phenotypes, both proportions increased to >95%. However, the total number of significant aseQTLs was smaller than the number of cis-eQTLs: the magnitude-based approach confirmed only 11.2–18.4% of cis-eQTLs and contributed <1% additional candidate signals relative to the number of cis-eQTLs, whereas the direction-based approach confirmed 39.4–52.3% of cis-eQTLs and added 0.4–9.2% additional signals. The lower power of ASE relative to cis-eQTL mapping is expected, because ASE requires informative heterozygous sites and accurate phasing and is therefore available only for a subset of genes and individuals, whereas cis-eQTL mapping leverages data across all individuals. In addition, the direction-based method showed greater power to validate cis-eQTLs and identify additional candidates than the magnitude-based method (3-4-fold increase in signals), while >91% of magnitude-based signals were also detected by the direction-based method (**Figure S9b**). We also observed the expected dependence on sample size: the number of aseQTLs increased with sample size for both methods and the fraction of recovered cis-eQTLs increased with sample size. For variant–gene pairs significant in both ASE and cis-eQTL analyses, effect sizes were highly concordant. The ASE effect size, expressed as the median log aFC across phased heterozygotes, closely mirrored the cis-eQTL slope estimate (Spearman’s ρ = 0.90–0.92 for the magnitude-based and 0.88–0.89 for the direction-based method; **Figure S9c-d**). Together, these results show that ASE provides strong orthogonal support for our cis-eQTL discoveries across GNs: the vast majority of detected aseQTLs recapitulate cis-eQTL signals with concordant directions of effects.

**Supplementary Figures**

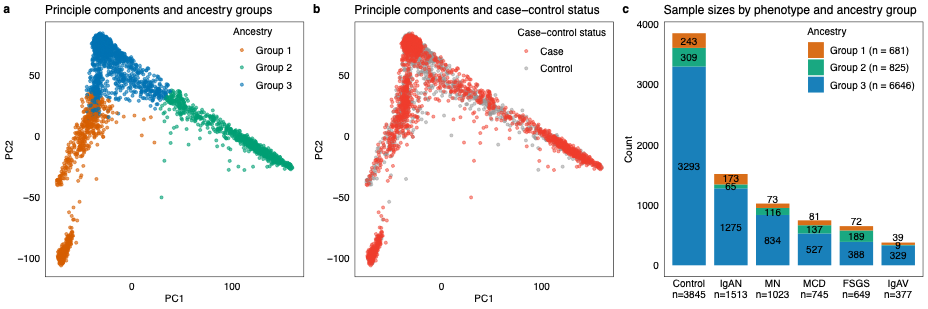

**Supplementary Figure 1. Principle components and genetic ancestry of participants with whole genome sequencing data.** (a) The cohort comprised 8,152 individuals and was stratified into three genetic ancestry groups: Group	1 (N = 825, orange), Group 2 (N = 681, green), and Group 3 (N = 6,646, blue). (b) Genetic population structure was concordant between cases representing patients across multiple glomerular disease subtypes (N = 4,307) and controls (N = 3,845). (c) Distribution of study participants across GN subtypes and ancestry groups.

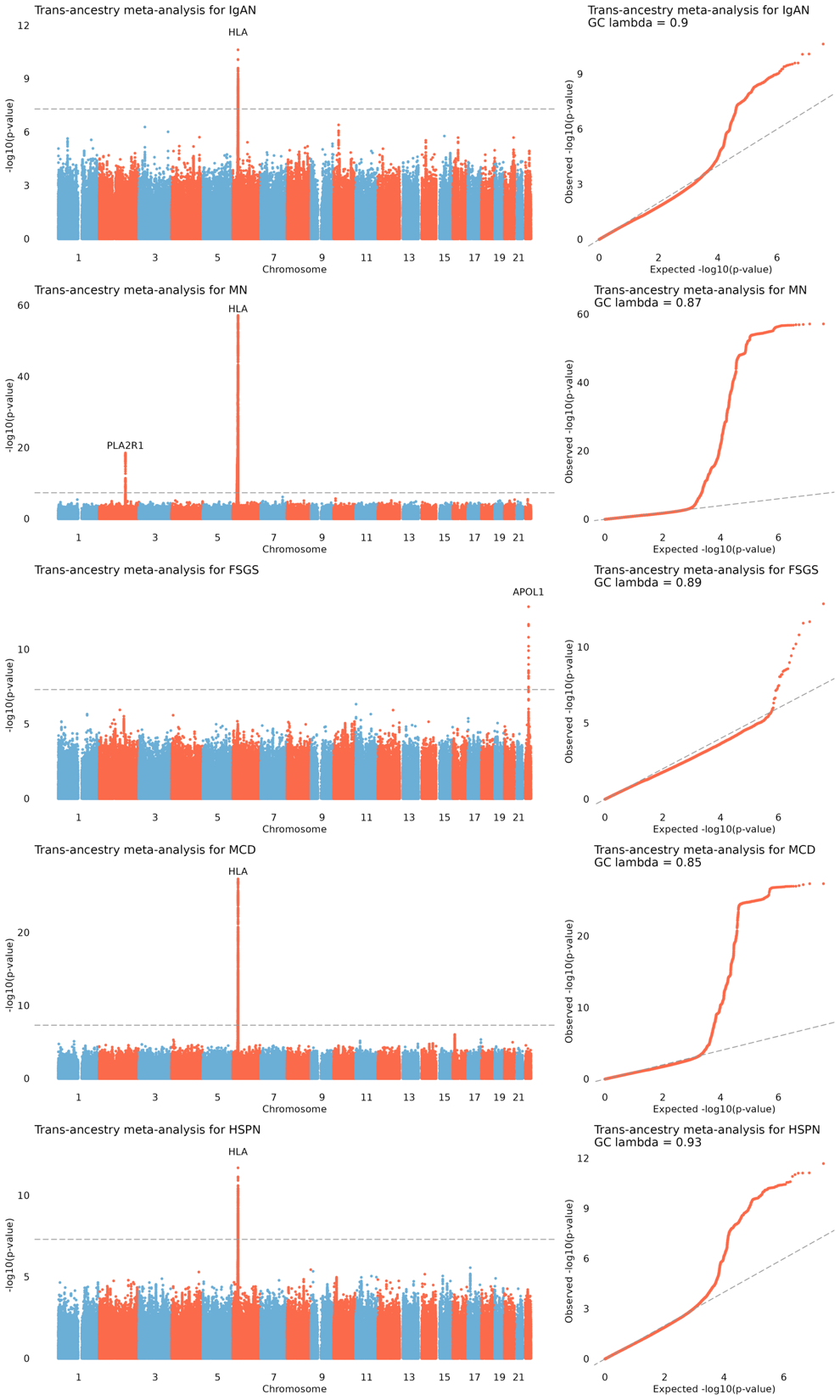

**Supplementary Figure 2. Manhattan and Quantile-quantile (qq) plots for common variant association testing.** (a) Trans-ancestry meta-analysis for IgA nephropathy (Ncase = 1505; Ncontrol = 3417). The overall genome inflation factor (lambda) was estimated at 0.9. (b) Trans-ancestry meta-analysis for membranous nephropathy (Ncase = 1022; Ncontrol = 3417). The overall genome inflation factor (lambda) was estimated at 0.87. (c) Trans-ancestry meta-analysis for Focal segmental glomerulosclerosis (Ncase = 645; Ncontrol = 3417). The overall genome inflation factor (lambda) was estimated at 0.89. (d) Trans-ancestry meta-analysis for minimal change disease (Ncase = 743; Ncontrol = 3417). The overall genome inflation factor (lambda) was estimated at 0.85. (e) Trans-ancestry meta-analysis for IgA vasculitis (Ncase = 377; Ncontrol = 3417). The overall genome inflation factor (lambda) was estimated at 0.93. In Manhattan plots, the dotted horizontal line indicates a genome-wide significance threshold (*α* = 5 × 10^−8^)

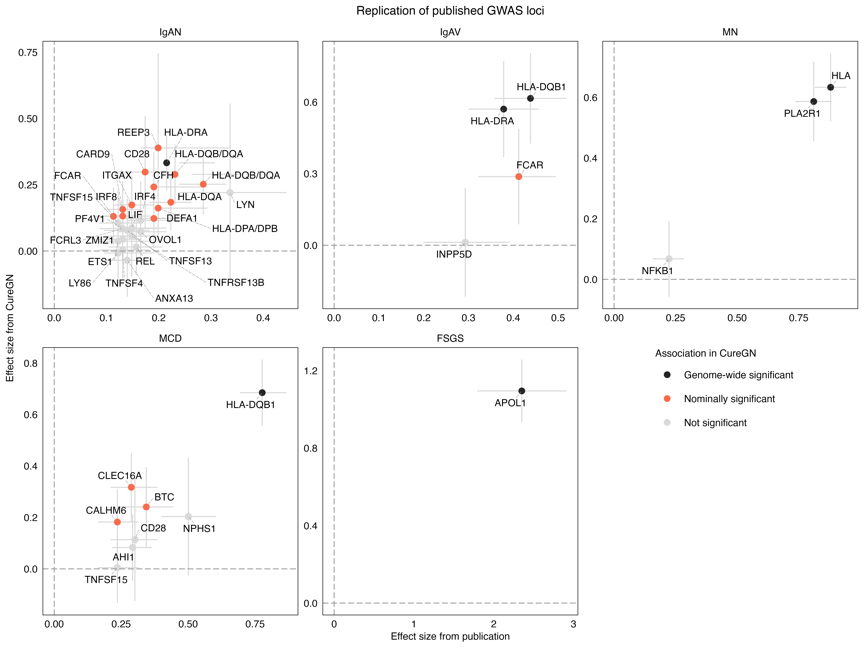

**Supplementary Figure 3.** **Replication of published GWAS loci for five glomerular disease subtypes in CureGN.** Each point represents a reported GWAS locus with effect size measure as beta = log(OR) from previously published GWAS (x-axis) and corresponding effect size estimated in the CureGN cohort (y-axis). Error bars indicate 95% confidence intervals for effect size estimates. Points are colored by significance level in CureGN: genome-wide significant (black), nominally significant (orange), and not significant (gray). Glomerular disease subtypes: IgA nephropathy (IgAN), IgA vasculitis (IgAV), membranous nephropathy (MN), minimal change disease (MCD), and focal segmental glomerulosclerosis (FSGS).

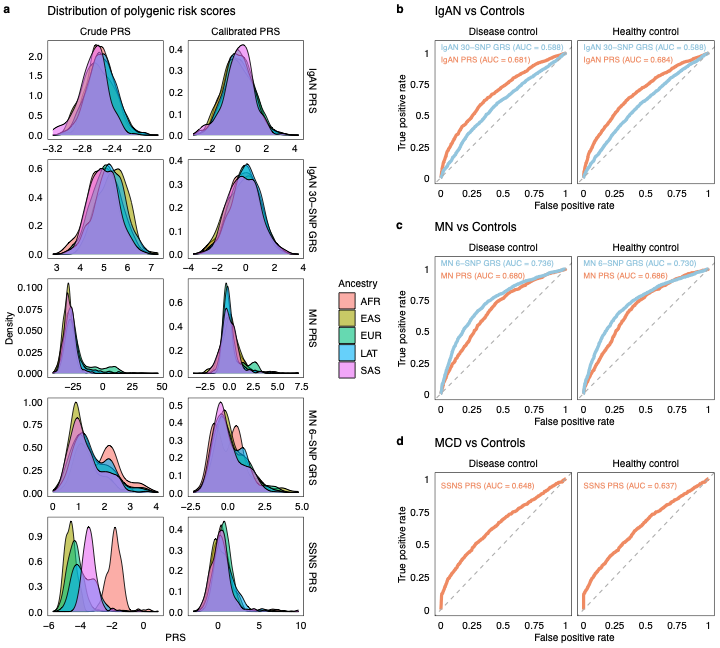

**Supplementary Figure 4. Genome-wide polygenic risk scores.** Distribution and predictive performance of polygenic risk scores across ancestries and disease comparisons. (a) Distribution of polygenic risk scores (IgAN PRS, MN PRS, and SSNS PRS) and genetic risk scores (IgAN 30-SNP GRS and MN 6-SNP GRS) across five ancestry groups (AFR, EAS, EUR, LAT, and SAS) before (crude) and after ancestry adjustment (calibrated). (b-d) Receiver operating characteristic (ROC) curves and area under the curve (AUC) values evaluating the predictive performance of PRS models for discriminating cases from disease controls and healthy controls. (b) IgAN PRS compared with 30-SNP GRS for IgAN versus controls; (c) MN PRS compared with MN 6-SNP GRS for MN versus controls; (d) SSNS PRS for MCD versus controls.

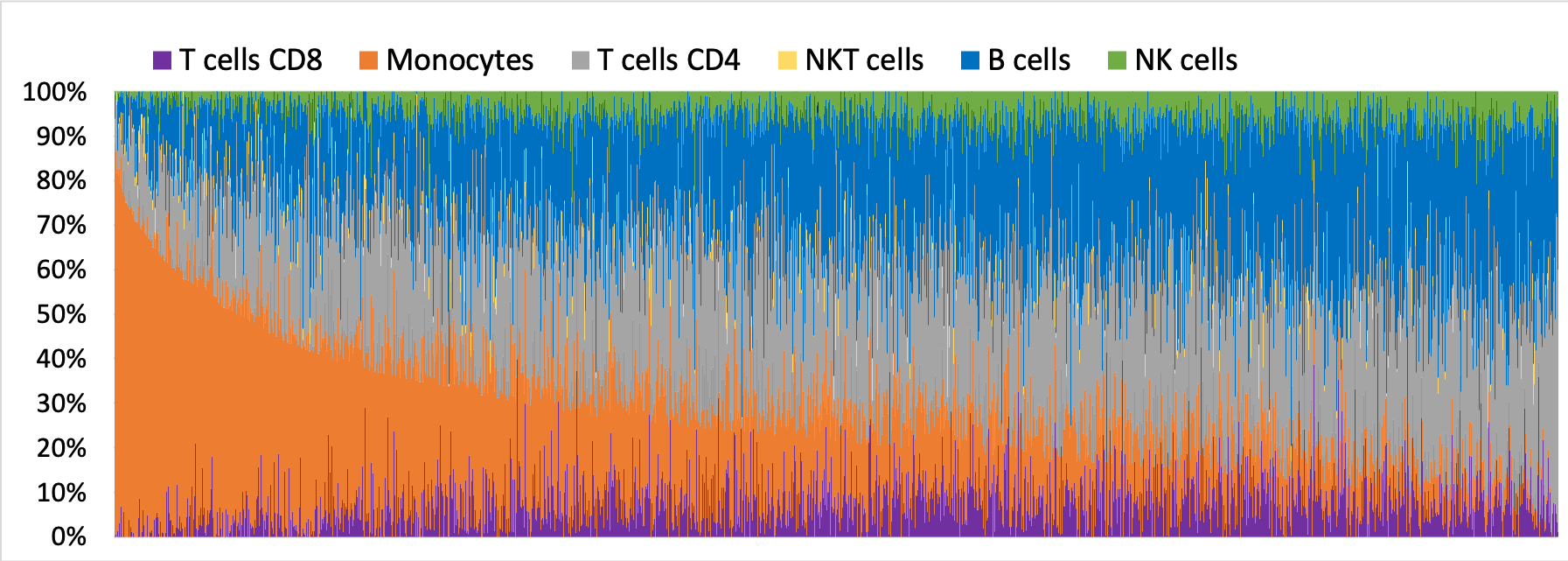

**Supplementary Figure 5. The distribution of cell type fractions across all 1,822 CureGN participants.** Bulk blood RNA-seq gene expression across individuals was deconvoluted to estimate contributions from six major blood cell types: B cells, monocytes, CD4⁺ T cells, CD8⁺ T cells, natural killer (NK) cells, and natural killer T (NKT) cells. Individuals were ordered by their monocyte proportions.

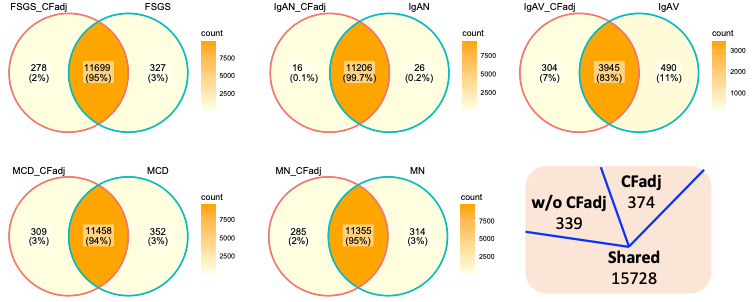

**Supplementary Figure 6. Context-specific eQTL mapping with and without adjustment for cell type proportions.** Comparison of eQTLs identified with and without adjustment for cell type fractions. Most eQTLs (86–97%) were shared between the two analyses. A small fraction (1–5%) were detected only after adjusting for cell type proportions. Conversely, 2–9% of eQTLs identified without adjustment lost significance after accounting for cell type composition, suggesting these signals may be influenced by variation in cell type proportions across individuals.

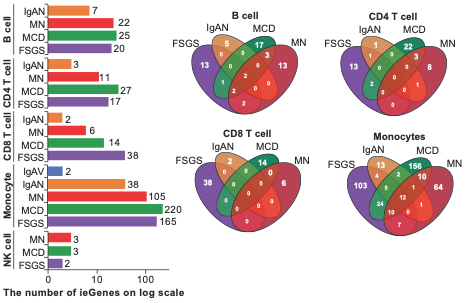

**Supplementary Figure 7. Interaction eQTL mapping with 5 major blood cell types.** The left bar plots show the number of ieGenes identified in B cells, CD4 T cells, CD8 T cells, monocytes, and NK cells across IgAN (orange), MN (red), MCD (green), FSGS (purple), and IgAV (monocytes only, blue). The counts are represented on a logarithmic scale and illustrate marked variation among disease subgroups, particularly for monocytes. The right Venn diagrams depict the overlap in ieGenes among disease groups for each immune cell type (B cell, CD4 T cell, CD8 T cell, monocyte), highlighting both shared and unique gene sets for IgAN, MN, MCD, and FSGS.

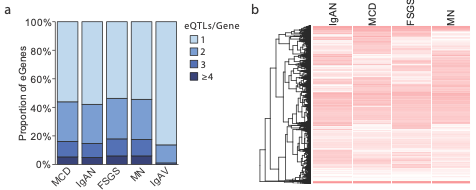

**Supplementary Figure 8. Independent eQTL signals and comparison of effect sizes across traits.** (a) Distribution of independent eQTL signals across different disease conditions. The y-axis is the proportion of identified eGenes for a given trait. (b) Hierarchical clustering of genome-wide eQTL effects across traits. Red indicates stronger genetic effects on gene expression and white indicate no genetic effects on gene expression.

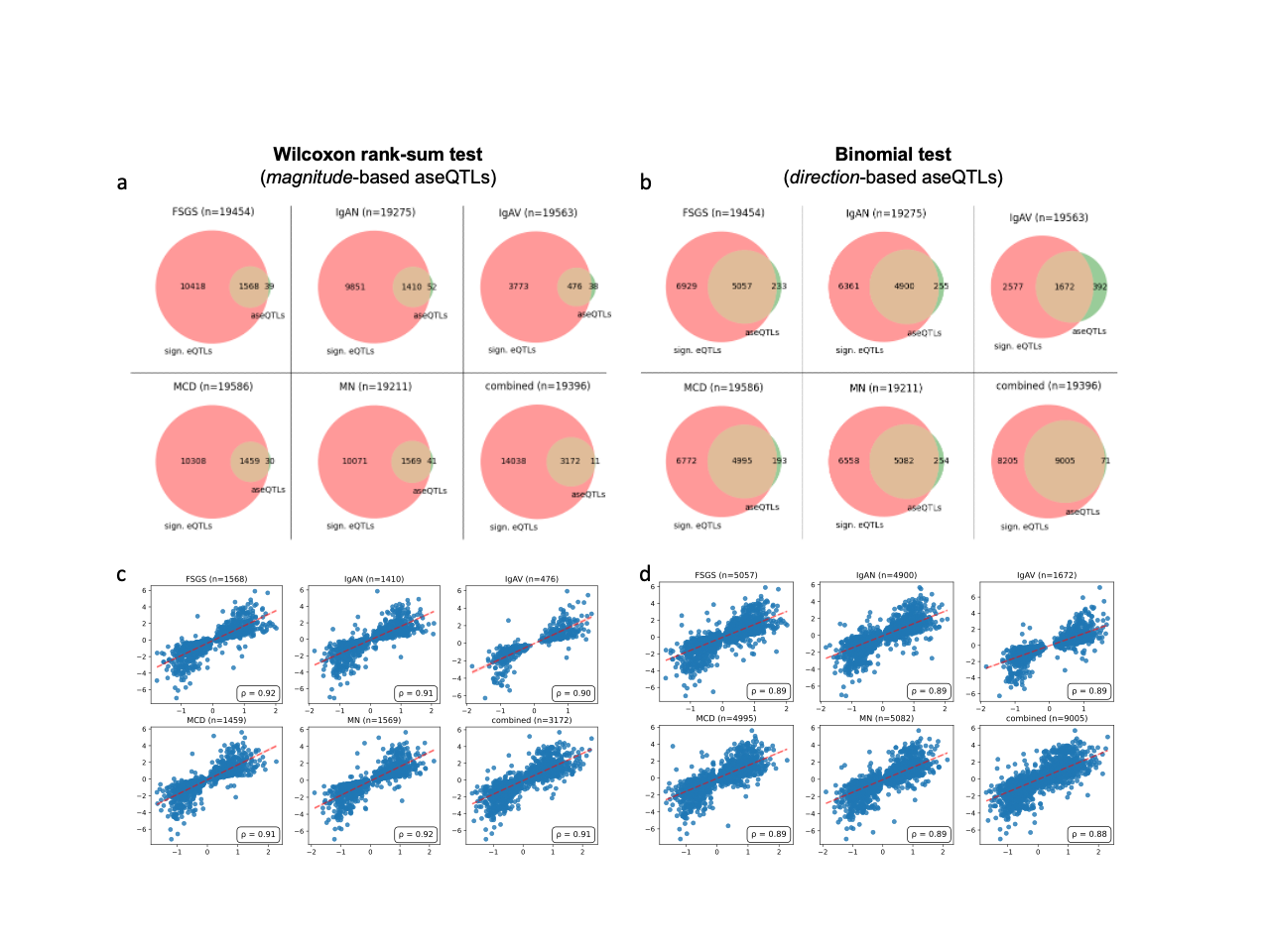

**Supplementary Figure 9. Comparison of significant cis-eQTLs and aseQTLs across different glomerular disorders, with correlation analyses.** **(a-b)** Venn diagrams illustrating the overlap between significant expression quantitative trait loci (cis-eQTLs, red) and allele-specific expression QTLs (aseQTLs, green) defined based on **(a)** wilcoxon rank sum test and **(b)** binomial test for each disease (FSGS, IgAN, IgAV, MCD, MN) and the combined cohort. Numbers within each section indicate the count of QTLs unique to each analysis and their overlap. **(c-d)** Scatterplots showing the correlation of effect size between overlapping cis-eQTLs and aseQTLs in each cohort: **(c)** for *magnitude*-based aseQTLs, **(d)** for *direction*-based aseQTLs. The linear regression line indicates the positive association between effect sizes identified by both methods. ρ denotes Spearman’s rank correlation coefficient.

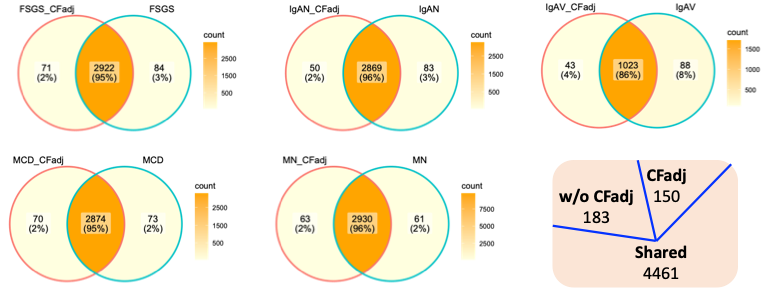

**Supplementary Figure 10. Context-specific sQTL mapping with and without adjustment for cell type proportions.** Similar to eQTL mapping, most eQTLs (86–96%) were shared between the two analyses. About 1–4% were detected only after adjusting for cell type proportions, and 2–8% of sQTLs identified without adjustment lost significance after accounting for cell type composition, suggesting these signals may be affected by variation in cell type proportions across individuals.

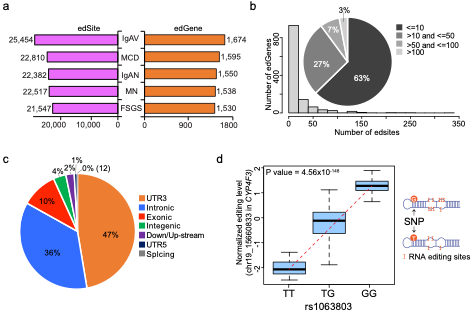
**Supplementary Figure 11. Context-specific edQTL mapping in glomerulonephropathy.** **(a)** Bar plots depicting the total number of RNA editing sites (edSites) and editing genes with significant QTL effects (edGenes, orange) in five glomerular diseases: IgA vasculitis (IgAV), minimal change disease (MCD), IgA nephropathy (IgAN), membranous nephropathy (MN), and focal segmental glomerulosclerosis (FSGS). **(b)** Distribution of RNA editing sites per gene. Left, histogram showing how many genes contain different numbers of editing sites. Right, pie chart summarizing the proportion of edGenes by categorized number of editing sites (≤10, >10 & ≤50, >50 & ≤100, >100). **(c)** The genomic annotation of RNA editing sites, including proportions in 3' untranslated regions (UTR3), intronic, exonic, intergenic, downstream/upstream, 5' untranslated regions (UTR5), and splicing sites. **(d)** Boxplot showing the top edQTL in the CYP4F3 gene stratified by rs1063803 genotype (TT, TG, GG). The Y-axis indicates normalized editing levels.

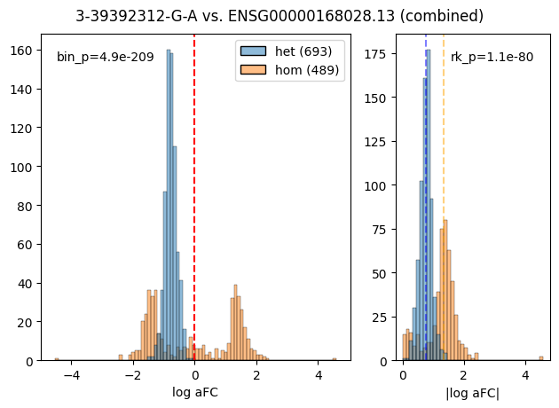

**Supplementary Figure 12.** The overdispersion of the log aFC in homozygous samples (due to lower overall detected expression in homozygous samples) causes the ranksum-statistics to be in the wrong direction – even though the two-sided p-value is very significant (1.1e-80), the corrected one-sided p-value is close to 1, while we see that the log aFCs in the heterozygous samples are clearly shifted to the left from the 0 line on the left panel, which is basically the definition of allele-specific expression, hence the binomial p-value is very significant in that case (p=4.9e-209). In less extreme cases, the homozygous distribution might be slightly less dispersed and then the two-sided rksum statistics for |log aFC| would also become insignificant, while the heterozygous samples would show the ASE effect via the binomial-based p-value.
